# Triple therapy prevention of Recurrent Intracerebral Disease EveNts Trial (TRIDENT)

**DOI:** 10.1101/2025.08.24.25334319

**Authors:** Laurent Billot, Sana Shan, Anthony Rodgers, Craig Anderson, Robin Bliss

## Abstract

The TRIDENT trial aims to determine the effectiveness and safety of a single pill combination of three low-dose components (telmisartan 20mg, amlodipine 2.5mg and indapamide 1.25mg) versus placebo on top of standard of care, in patients with a history of stroke due to intracerebral hemorrhage. It is designed as an international, multicenter, double-blind, placebo-controlled, parallel-group, randomized controlled trial. The primary outcome is time from randomization to first occurrence of recurrent stroke, whether intracerebral hemorrhage, ischemic or undifferentiated.

This statistical analysis plan pre-specifies the method of analysis for key outcomes and variables collected in the trial. The primary analysis will consist of a Cox proportional hazard adjusted for stratification variables. The analysis plan also includes planned sensitivity analyses including covariate adjustments and subgroup analyses.

An investigator initiated and conducted, multicenter, international, double-blind, placebo-controlled, parallel-group, randomized controlled trial to determine the effectiveness of more intensive blood pressure control provided by a fixed low-dose of blood pressure lowering agents as a single pill combination ‘Triple Pill’ strategy on top of standard of care, on the time to first occurrence of recurrent stroke in patients with a history of stroke due to intracerebral hemorrhage.

## 1 Administrative information

### 1.1 Study identifiers

- Protocol Number: GI-AU-NMH-2016-01, Version 6.0, Date 6 December 2022
- ClinicalTrials.gov register Identifier: NCT02699645

### 1.2 Revision history

| Version | Date | Details |
| --- | --- | --- |
| 1.0 | 1 April 2025 | Initial draft prior to database lock – sent to FDA for review |
| 2.0 | 16 June 2025 | Updated draft prior to database lock – sent to FDA for review following receipt of initial FDA feedback on v1.0.<br>Updates include a reordering of secondary and exploratory endpoints, clarification of type I error methods, and clarification of analysis methods. See Section 7 for further details in Revision History: Summary of Changes. |
| 3.0 | 22 August 2025 | Updated draft prior to database lock<br>Inclusion of protocol deviations definitions<br>Inclusion of analysis of MRI data<br>Inclusion of listing of planned publications<br>Inclusion of figure and table shells<br>Other minor changes |

### 1.3 Contributors to the statistical analysis plan

#### 1.3.1 Roles and responsibilities

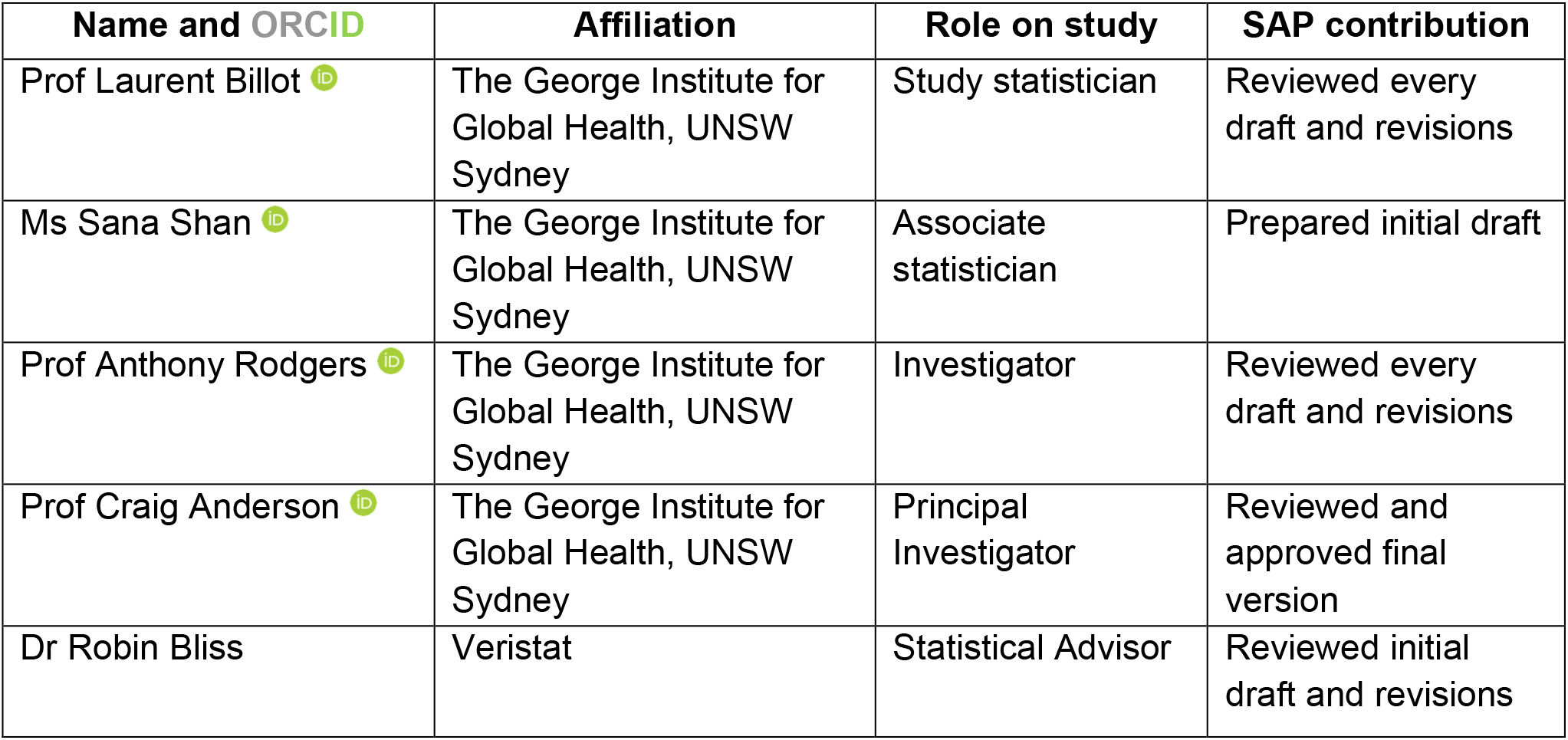

1.3 Contributors to the statistical analysis plan 1.3.1 Roles and responsibilities
| Name and ORCID | Affiliation | Role on study | SAP contribution |
| --- | --- | --- | --- |
| Prof Laurent Billot | The George Institute for Global Health, UNSW Sydney | Study statistician | Reviewed every draft and revisions |
| Ms Sana Shan | The George Institute for Global Health, UNSW Sydney | Associate statistician | Prepared initial draft |
| Prof Anthony Rodgers | The George Institute for Global Health, UNSW Sydney | Investigator | Reviewed every draft and revisions |
| Prof Craig Anderson | The George Institute for Global Health, UNSW Sydney | Principal Investigator | Reviewed and approved final version |
| Dr Robin Bliss | Veristat | Statistical Advisor | Reviewed initial draft and revisions |

#### 1.3.2 Approvals

The undersigned have reviewed this plan and approve it as final. They find it to be consistent with the requirements of the protocol and/or modifications to the study procedures as determined by the Steering Committee, to clarify the overall intention of the statistical analyses, and in response to FDA feedback (May and August, 2025). They also find it to be compliant with ICH-E9 principles and in particular, confirm that this analysis plan was developed in a completely blinded manner (i.e. without knowledge of the effect of the intervention[s] being assessed).

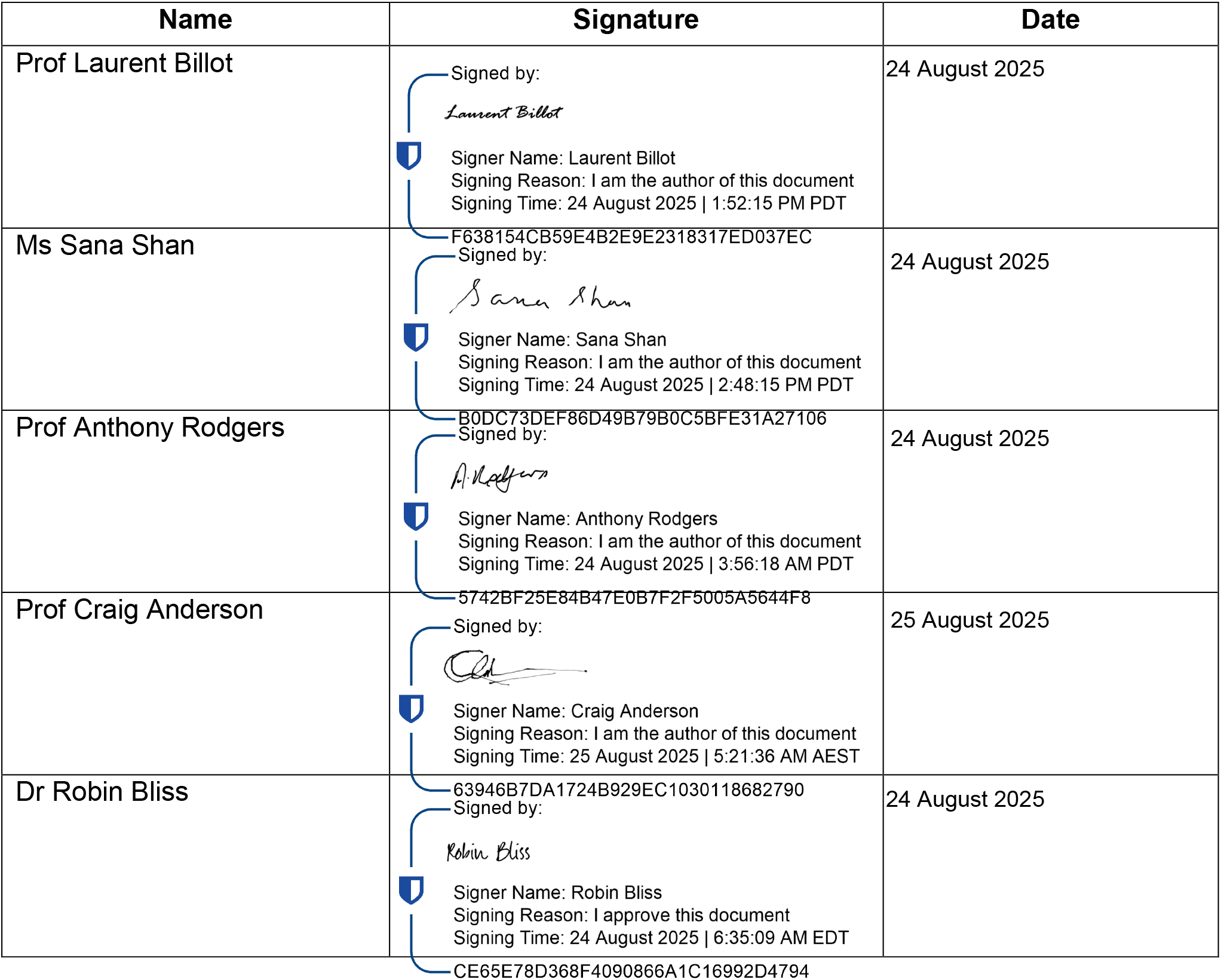

## 2 Introduction

### 2.1 Study synopsis

TRIDENT is an international, multicenter, investigator-initiated and conducted, double-blind, placebo-controlled, parallel-group, randomized controlled trial. The trial aims is to determine the effectiveness and safety of more intensive, long-term, blood pressure (BP) lowering provided by a single pill combination (SPC) of three low-dose components (telmisartan 20mg, amlodipine 2.5mg and indapamide 1.25mg) vs placebo on top of standard of care, on the time to first occurrence of recurrent stroke in patients with a history of stroke due to intracerebral hemorrhage (ICH). The full protocol was published in January 2022,^1^ and the last visit for last patient was undertaken in July 2025.

### 2.2 Study population

The study enrolled participants with a history of primary ICH from 12 countries. The double-blind treatment period was preceded by a 2-week, single-blind, active run-in phase in which all participants received the active intervention (telmisartan 20mg, amlodipine 2.5mg and indapamide 1.25mg). This active run-in aimed to ensure participants who were randomized were able to tolerate the treatment regimen and procedures and thus increase the likelihood that there would be high adherence to the follow-up schedule over several years.

#### 2.2.1 Inclusion Criteria

- Adults (≥18 years) with a history of primary ICH that was confirmed by imaging (copy of the brain imaging report was uploaded into the database, labelled with participant identification [ID] after removal of any personal identifiers).
- Clinically stable, as judged by investigator.
- An average of two resting systolic blood pressure (SBP) levels measured 5 minutes apart in the range 130-160 mmHg recorded in a seated position. Patients with higher SBP could still be included if it was considered by the site investigator that their management would be consistent with local standards of clinical practice.
- Geographical proximity to the recruiting hospital and/or follow-up medical clinic site to allow ready access for in-person assessments during follow-up.
- No clear contraindication or indication to any of the individual components of the Investigational Medical Product (IMP).
- Provision of written informed consent

#### 2.2.2 Exclusion Criteria

- Taking an angiotensin converting enzyme inhibitor (ACE-I) that could not be switched to any of the following alternatives: an angiotensin receptor blocker (ARB) (e.g. telmisartan 20 or 40 mg), a calcium channel blocker (CCB) (e.g. amlodipine 2.5 or 5 mg), or a thiazide (TZ)-like diuretic (e.g. indapamide 1.25 mg); an equivalent class (ARB, CCB or TZ); or a beta-blocker (BB).
- Contraindication to any component of the IMP in the context of currently prescribed BP lowering treatment.
- Unable to complete the study procedures and/or follow-up.
- Females of child-bearing age and capability, who are pregnant or breast-feeding, or those of child-bearing age and capability who are not using adequate birth control.
- Clinically significant hyperkalemia and/or hyponatremia, in the opinion of the responsible physician.
- Estimated glomerular filtration rate (eGFR) <30 mL/min/1.73m^2^.
- Severe hepatic impairment (alanine aminotransferase [ALT] or aspartate aminotransferase [AST] that is >3x the upper limit of normal [ULN]).
- Any other condition that in the opinion of the responsible site investigator renders the patient unsuitable for the study (e.g., severe disability such as scores of 4 or 5 on the modified Rankin scale [mRS], or significant memory or behavioural disorder)

### 2.3 Study interventions

#### 2.3.1 Run-in phase

Participants who were eligible for the run-in treatment, received single-blind (only participants remained blind) active intervention containing telmisartan 20 mg, amlodipine 2.5 mg and indapamide 1.25 mg, for a minimum period of 2 weeks to ensure that they who were able to tolerate the treatment regimen and procedures. A second 2-week run-in phase (repeat run-in) was an option for investigators, if the first period was interrupted, problematic or for any other reason due to unforeseen or unavoidable circumstances. Thus, all participants could have a maximum of 4 weeks to complete the run-in phase.

Successful completion of the run-in period was determined by participants being able to achieve at least 80% adherence to run-in medication and tolerability to the IMP and protocol.

Participants with eGFR <30 mL/min/1.73m^2^, or significant liver disease, hyperkalemia or hyponatremia, following the run-in phase were ineligible to proceed to randomization.

#### 2.3.2 Randomization

After successful completion of the run-in phase, participants were randomized via a secure web-based database, stratified to ensure a balance in the key prognostic factors: country of recruitment, age (<65 vs. ≥65 years) and mean of 2 recorded baseline SBP measurements (<140 vs ≥140 mmHg). All participants and study team members were kept blind to the randomization allocation; only the unblinded study statistician and manufacturer/supplier of the IMP were aware of the treatment allocation.

#### 2.3.3 Study treatment

Each participant received either active treatment (telmisartan 20 mg, amlodipine 2.5 mg, and indapamide 1.25 mg) or placebo respectively. The IMP that was first used in the study was an over-encapsulation of individual, standard, regulatory-approved, medications or an over-encapsulation of 3 matching placebos (from 28 September 2017). The IMP was switched to a bespoke SPC called GMRx2, developed by George Medicines Pty Ltd, with a matching placebo (from June 2022). In both situations, the study medication had identical packaging, labelling and administration scheduling.

### 2.4 Outcomes

#### 2.4.1 Primary outcome

- Time to first occurrence of recurrent stroke, whether ICH, ischemic or undifferentiated.

#### 2.4.2 Secondary efficacy outcomes

- Time to first occurrence of MACE, a composite of major adverse cardiovascular (CV) events (CV death, non-fatal myocardial infarction [MI], or non-fatal stroke)
- Time to CV mortality
- Hypertension control, defined as SBP <130 mmHg at 6 months follow-up

#### 2.4.3 Safety outcomes

- Withdrawal of treatment due to adverse event
- Serious adverse events (SAEs) according to standard definitions
- Adverse events of special interest (AESIs) – headache, falls, pedal edema, hypo/hyperkalemia and hyponatremia

#### 2.4.4 Other outcomes

The following outcomes will be regarded as exploratory.

##### 2.4.4.1 Other CV and mortality outcomes

- Time to first recurrence of ICH
- Time to first occurrence of ischemic stroke
- Time to first occurrence of fatal stroke
- Time to first occurrence of stroke of undifferentiated origin
- Time to nonfatal MI
- Time to all-cause mortality

##### 2.4.4.2 Cognitive outcomes

- Dementia-free survival, defined as a diagnosis of probable all-cause dementia at the end of follow-up, was defined according to the fifth Diagnostic and Statistical Manual of Mental Disorders (DSM V) criteria assessed in survivors at the end of the study or death due to any cause.
- Mild cognitive impairment (MCI), again according to DSM V criteria.
- Composite of dementia and MCI diagnosis.
- Cognitive decline, defined as a decline in global cognitive function based upon scores on the Montreal Cognitive Assessment (MoCA)^**2**^ assessed by site investigators at regular intervals during follow-up. Changes in the domain-specific aspects of cognitive function, such as learning and memory, complex attention, executive functioning, language, and visuospatial skills, will also be determined.

##### 2.4.4.3 Disability outcomes

- Disability-free survival.
- Physical functioning, according to scores on the simplified mRS (smRS).^4^
- Health-related quality of life, using the EuroQoL Group 5-Dimension 3-level self-report questionnaire

##### 2.4.4.4 Other BP outcomes

- Hypertension control throughout follow-up, defined as mean of all post-randomization measures SBP <130 mmHg.
- Hypertension control at end of study, defined as SBP <130mmHg at final follow-up visit.
- Change in SBP from randomization.
- Change in DBP from randomization.
- Time at target (SBP <130 mmHg) throughout follow-up.

##### 2.4.4.5 Other exploratory outcomes

Other exploratory outcomes include:

- In English-speaking participants, the Brief Memory and Executive Test (BMET)^3^ is also used as a more sensitive measure of executive functioning/processing speed administered in the context of cerebral small vessel disease (CSVD).
- In participants who survive to the end of follow-up, the degree and specific markers of CSVD and total brain volume will be assessed on brain MRI.
- Medication adherence (self-report and pill counts).
- Other MRI-based outcome measures.

## 3 Definitions of endpoints

### 3.1 Stroke

Stroke is defined as the sudden onset of focal neurological impairment or deficit with symptoms lasting for more than 24 hours, or reversing within 24 hours if this is accompanied by an acute ischemic lesion within an appropriate vascular territory being evident of brain imaging, generally an MRI being more sensitive than CT. Symptoms that resolve completely within 24 hours are considered to be a transient ischemic attack (TIA) and are not included as a stroke endpoint.

A non-fatal stroke event *does not result in death within 28 days from onset*. Any recurrence or exacerbation of the condition within 28 days is considered part of the original episode, except when it results in a different vascular territory. Any new neurological symptom(s) that occur beyond 28 days is considered as a separate event.

Stroke events are classified as ischemic, hemorrhagic (intracerebral or subarachnoid), or unknown etiology, based on supporting source documents. Ischemic strokes are classified as definite or probable: large artery atherosclerosis (>50% stenosis of extra or intra cerebral vessels); cardio-embolism (clear cardiac source of embolism such as atrial fibrillation with large vessel, peripheral or multiple territory cerebral infarction); small vessel related associated with lacunar syndrome; or other/uncertain in relation to mixed pathology or insufficient information to provide diagnosis. Definite or probable ICH is categorized as deep (cerebral hemisphere, brainstem and cerebellum), cortical (lobar) or uncertain. Other pathological categories of stroke are subarachnoid hemorrhage and stroke of undetermined etiology.

All stroke events will be independently adjudicated by an expert panel, blind to allocation.

### 3.2 Myocardial infarction

The diagnosis of MI requires the following criteria to be met: detection of a rise or fall of a cardiac biomarker (preferably troponin) with at least one value above the 99th percentile of the upper reference limit (URL) together with evidence of myocardial ischemia with at least one of the following: ischemic signs or symptoms (i.e., chest, arm, neck, or jaw discomfort; shortness of breath, pulmonary edema); pathologic Q waves present in any two contiguous ECG leads that are >30 milliseconds; new or presumed ECG changes indicative of ischemia, either ST-T wave abnormalities in ≥2 contiguous ECG leads (i.e. ST segment elevation >2 mm in leads V1, V2, or V3, or >1 mm in the other leads), or ST segment depression (>1 mm) or symmetric inversion of T waves >1 mm; OR new LBBB; imaging evidence of loss of viable myocardium that is presumed to be new and, in a pattern, consistent with an ischemic etiology (i.e. new cardiac wall motion abnormality on echocardiography or new fixed defect on radionuclide imaging or new myocardial scarring or edema on cardiac MRI); identification of intracoronary thrombus on angiography or autopsy

MI is further classified as: ST elevation MI, defined as new ST elevation of 1 mm in limb leads or 2 mm in precordial leads and typical symptoms (e.g. chest pain); silent Q-wave MI associated with the development of new Q waves of duration ≥0.03 duration in ≥2 ECG leads in the same lead group and in the absence of left bundle branch block (LBBB) or ventricular pacing (as per the Minnesota Code Classification System; non ST elevation MI defined as the presence of new and persistent (>24 hours) ST changes or T wave changes defined as ST depression of 3mm or T wave inversion of 3 mm on the ECG with significant cardiac enzyme/marker elevation (as above) and/or typical chest pain; MI without significant ECG changes in a patient with characteristic symptoms plus significant elevation of cardiac enzymes/markers; peri-procedural MI in relation to a specific cardiac percutaneous procedure (within 7 days) or coronary artery bypass graft (within 30 days) or non-CV surgical interventions (within 30 days).

All MI events are independently adjudicated by an expert panel, blind to allocation.

### 3.3 Cause of death

Definition of primary cause of death – the event which immediately preceded death. All deaths will be independently adjudicated by an expert panel, blind to allocation.

### 3.4 CV death

Death from any CV cause is defined as any death in which the primary or the underlying cause of death is due to a disease of the circulatory system or unexpected sudden death. The death was categorised as stroke, MI, or sudden presumed to be due to ischemic CV disease, occurring within 24 hours from onset of symptoms without confirmation of CV cause, and without clinical or autopsy evidence of etiology, or un-witnessed death where the body of the deceased was found dead for which no cause can be discovered. In cases where it is not possible to determine the cause of death as definitely CV-related, the event will be classified as non-CV death.

### 3.5 Dementia

The assessment of cognitive function in all participants was undertaken at regular intervals during follow-up. This includes an in-person cognitive screening assessment that is administered at baseline, 6 months, and 18 months, and then every 12 months until the end of follow-up. This assessment involved two tests of global cognitive function, first on the Montreal Cognitive Assessment (MoCA; range 0-30)^2^ and the other on the BMET^3^ (range 0-16) in English-speaking participants as a more sensitive measure to detect problems of frontal executive cognitive function.

The assessment for a diagnosis of dementia or MCI is undertaken via a 3-step process, either at the penultimate or final visit of participants, undertaken between late 2024 and early-mid 2025. Any participant who scored <26 or had a decline of ≥3 points in the previous 12 months on the MoCA is deemed to be ‘screen positive’, which requires them to have a further assessment by a trained researcher, either a neuropsychologist (in Sri Lanka) or neurologist (other regions). This person undertook a further assessment of cognitive function through in-person administration of the Quick Dementia Rating Scale (QDRS)^5^ and the 12-item Patient Health Questionnaire (PHQ-12)^6^ to the participant; and in-person administration of the short Informant Questionnaire for Cognitive Decline in the Elderly (IQCODE)^7^ and the completion of several short questions to a reliable informant:

1. Since the person sustained their stroke, do you feel that their memory and thinking skills have declined?
2. Since the person sustained their stroke, do you feel that the person has decline in his/her functioning due to difficulties with memory and thinking?
3. How would you characterize the person’s decline in their memory and thinking: stable since the stroke or stepwise progressive since the stroke
4. Has the patient ever been diagnosed with dementia?
5. Has the patient displayed any signs of cognitive impairment?

Trained expert neurology-certified adjudicators classified participants into 1 of 4 primary categories: no cognitive impairment, uncertain cognitive impairment, MCI, or probable dementia. Each case is reviewed independently by at least 2 adjudicators using standardized diagnostic criteria for probable dementia and MCI. Agreement by at least 2 adjudicators is final; disagreements required a third or rarely a forth adjudicator. No subclassification of probable dementia is made. Further details of the adjudication process can be found in the trial protocol.

#### 3.5.1 Diagnosis of dementia and MCI

Based on DSM V criteria, the diagnosis of probable dementia is present if there was evidence of *significant cognitive impairment that is sufficient to interfere with independence* (i.e. requiring assistance with instrumental activities of daily living), defined by each of the following items being present:

1. impairment in ≥1 cognitive domain on the QDRS, and for the overall score to be ≥7;
2. the IQCODE score is ≥4;
3. the informant notes a decline in cognitive function in a stepwise progression in memory and thinking since the stroke;
4. there is no evidence of major depression (PHQ-12 score <15) or significant another neurological or mental disorder.

The diagnosis of MCI is made if there is evidence of *modest cognitive decline* in ≥1 domain of cognitive function *without loss of independent function* (e.g. instrumental activities of daily living tasks are preserved), although they may still require greater effort, prompting, compensatory strategies, or accommodation, defined by each of the following being present:

1. impairment in ≥1 cognitive domain on the QDRS, with overall score of ≥3 and <7;
2. the IQCODE score is <4;
3. the informant notes the participant has normal or stable cognitive function since the stroke;
4. there is no evidence of major depression (PHQ-12 score <15) or another neurological or mental disorder

Normal cognitive function was defined as scores <3 and <4 on the QRDS and IQCODE, respectively.

## 4 Analysis principles

### 4.1 Sample size

The study was originally designed to recruit 3782 participants (1891 per treatment group) to have approximately 230 participants with a confirmed stroke outcome. This estimate was based on an annual control event rate of 2.5% and a hazard ratio (HR) of 35%. However, the sample was revised in early 2020^**1**^ after challenges were experienced in recruitment and with publication of the results of the Japanese Recurrent Stroke Prevention Clinical Outcome (RESPECT) trial.^**8**^ RESPECT suggested a large benefit of intensive BP lowering for the prevention of ICH but it was in a relatively small population. However, a large treatment effect was also seen in the Perindopril Protection against Recurrent Stroke Study (PROGRESS) trial, which was a key rationale for TRIDENT. The sample size was then revised with blinding being maintained by the Steering Committee without any knowledge of the treatment effect in the study. Overall, 1500 participants was estimated to provide 90% power (2-sided type-I error rate of 5%) to detect a HR of 0.5 (50% reduction) based on a total primary event rate of 12% over a mean of 3 years of follow-up (4% per year); this corresponds to a total of 100 patients with a primary outcome event. The calculation accounted for 5% lost to follow-up and 10% crossover (5% drop-in and 5% drop-out). None of the deliberations on revised sample size were made from the unblinded interim data, and the Steering Committee and study team remained blinded throughout the study.

### 4.2 Software

Analyses will be conducted primarily using SAS Enterprise Guide (version 8.3 or above) and R (version 4.0 or above).

### 4.3 Data and Safety Monitoring Board

An independent Data Safety Monitoring Board (DSMB) was convened with the responsibility of confidentially reviewing unblinded data relating to treatment efficacy, participant safety and quality of trial conduct, in accordance with the DSMB Charter put in place at the start of the study. The DSMB provided recommendations to the Study Steering Committee for the continuation of the study. The Steering Committee and all related study staff remained blinded to study results.

Two formal, pre-specified, unblinded, interim analyses were planned but only one was undertaken in April 2022. The unblinded results were reported by a designated unblinded statistician, to the independent DSMB, as was outlined in the DSMB charter that was finalized at the start of the study. The Haybittle-Peto stopping rule was applied to account for the potential for early stopping for efficacy.^9^ The DSMB recommended to the Study Steering Committee that the study proceed without modification throughout the study period. Given the conservative early stopping rule and the negligible amount of type-I error rate spent at the interim analysis, the significance threshold will remain at 5% (2-sided) for the final analysis. Only the designated unblinded statistician and the independent DSMB had access to unblinded data. All individuals involved in the design and conduct of the trial remained blinded to the study results and were not provided with any interim data by treatment group for review.

### 4.4 Multiplicity adjustment

All tests are to be two-sided with a nominal level set at 5%. Given the negligible amount of alpha spent at the interim analyses using Haybittle-Peto boundaries, analyses of the primary endpoint (time to first occurrence of recurrent stroke) will be performed with an unadjusted type I error rate. Components of the primary endpoint including (1) recurrent ICH, (2) ischemic (and undifferentiated) stroke and (3) fatal stroke, will be summarized with descriptive statistics without multiplicity adjustment applied. Each component will be analyzed regardless of statistical significance on the primary composite endpoint, but with only point estimates and 95% confidence intervals (CI) reported without significance tests.

For the 3 secondary efficacy endpoints (Section 2.4.2), we will control the family-wise error rate by applying a sequential Holm-Sidak correction.^10^ Briefly, the Holm-Sidak approach consists of ordering all p-values from smallest to largest, and then comparing them to an adjusted level of significance calculated as 1-(1-0.05)^1/C^, where C indicates the number of comparisons that remain. In the case of 3 secondary endpoints, the smallest p value would be compared to 1-(1-0.05)^1/3^, the second p value to 1-(1-0.05)^1/2^, with the last one being compared to 1-(1-0.05) (i.e. 0.05). The sequential testing procedure stops as soon as a p value fails to reach the corrected significance level. In case of sensitivity analyses (i.e. further adjustments), this will apply only to the primary analysis.

No multiplicity adjustment will be applied to other endpoints including safety endpoints, exploratory endpoints and sensitivity analyses.^3^

### 4.5 Data sets analyzed

All analyses will be conducted on an intention-to-treat (ITT) principle. The ITT population is all patients randomized regardless of whether they receive study treatment post-randomization according to the protocol.

### 4.6 Handling of the ‘end of study’ visit

Given study participants enter the study at different times and are followed for various durations, the end of study (EOS) visit will occur at different times for different participants. For longitudinal and descriptive analyses by visit, EOS visits will be re-allocated according to the planned visit schedule (i.e. every 6 or 12 months, depending on the nature of the data being collected). The allocation, based on the number of days between the date of randomization and the EOS visit, will be done as follows:

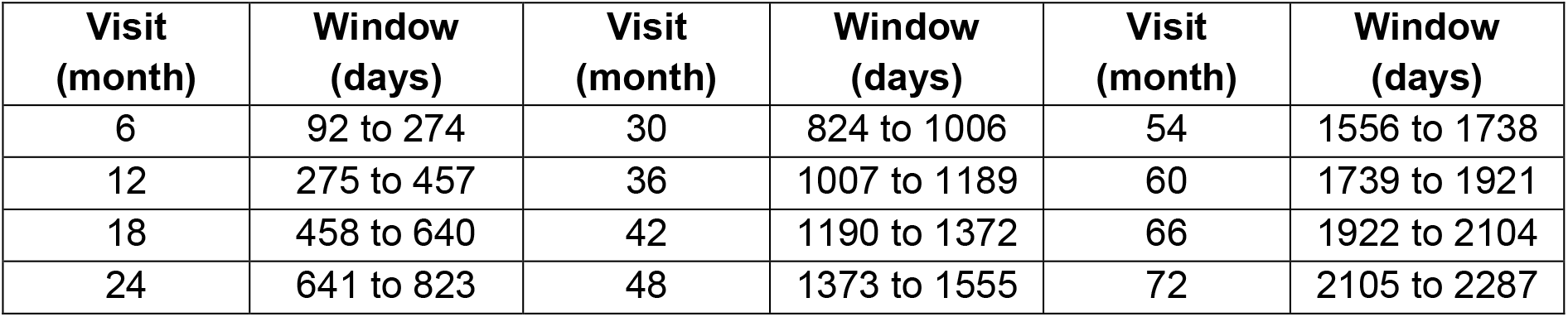

In case of a reallocated EOS visit falling within a window already containing a scheduled visit, we will retain the value from the previous scheduled visit, unless it is missing.

### 4.7 Missing Data

In general, summary statistics will be presented by visit, including all those with measurements at each respective visit without imputation for missing values.

Time to event endpoints will be analyzed using all available information with censoring rules as defined in the respective analysis sections.

For control of hypertension, all available information will be included in the analysis. Participants with missing hypertension assessments at 6 months follow-up will be imputed as having uncontrolled hypertension for the purpose of the secondary endpoint analysis.

For exploratory endpoints measured only at the last follow-up visit (i.e., dementia and MCI), analyses will only include participants with measurements, as defined in the respective sections. Those without an end of follow-up assessment recorded will be excluded from analysis. Binomial regression models with offset terms will be implemented to account for the variable time of follow-up. As a sensitivity analysis, participants missing the end of follow-up assessment of dementia will be imputed as a dementia case in a worst-case scenario imputation. Similarly, participants missing the end of follow-up assessment of MCI will be imputed as MCI cases.

Hypertension control at the end of follow-up will be analyzed using all available information. A binomial regression where “success” is defined as hypertension control at the last recorded visit will be performed with an offset term to account for variable follow-up time.

In general, exploratory endpoint analysis, including cross-sectional and longitudinal models will be performed using all available data without imputation.

## 5 Planned analyses

### 5.1 Subject disposition

The flow of patients through the trial will be displayed in a Consolidated Standards of Reporting Trials (CONSORT)^11^ diagram (see Figure 1). The report will include the following: the number of screened patients who met study inclusion criteria and the number of patients who were included; and reasons for exclusion of non-included patients. This will include a detailed analysis of the counts and reasons for patients continuing after the single blind active run-in.

### 5.2 Patient characteristics and baseline comparisons

Baseline characteristics will be summarized by treatment groups. Discrete variables will be summarized by frequencies and percentages. Percentages will be calculated according to the number of patients for whom data are available. Continuous variables will be summarized using mean and SD, and median and interquartile range (Q1-Q3). Baseline measures for all patients will be tabulated for the variables listed below:

- Demographics and socio-economic status: includes age (years), sex, ethnicity, level of education, employment status, income, and household information
- Anthropometrics: weight (kg), height (cm) and BMI (kg/m^2^)
- Use of current BP lowering medication(s)
- Medical history, including hypertension, cardiac disease, and atrial fibrillation
- Lifestyle factors: smoking, alcohol, caffeinated drinks, and dietary intake
- BP home monitoring
- Physical function (smRS)
- Cognitive assessments: MoCA, BMET
- EQ-5D-3L assessment
- Clinical data (last value prior to randomization): glucose, total cholesterol, LDL, HDL, triglycerides, creatinine, sodium, potassium, blood urea nitrogen, eGFR, AST, ALT

### 5.3 Compliance and protocol deviations

Compliance with the randomized intervention will be summarized using the following variables obtained at each follow-up visit:

- Number of missed doses of IMP in the last week as self-reported by the participant.
- Number of capsules/pills remaining from returned medication.

Adherence will be determined primarily by using self-reported participant missed doses. Acceptable adherence will be defined as missing no more than 1 day out of 7 days in the week preceding the visit for 80% or more of follow-up visits. As a sensitivity analysis, the number of the pills remaining from returned medicine count will be used to confirm the participant’s self-reported missed dose data, defining adherence as returning less than 20% of the dispensed capsules.

Number of days with missed doses and number of capsules returned will be summarized as means, SD, median, and quartiles by visit. Adherence at each visit will be summarized as counts and proportions, and a simple unweighted average calculated for each participant across all follow-up visits. No formal models or tests will be performed.

Protocol deviations (PDs) will be categorized as either minor or major based on their impact on the trial’s integrity, the quality and completeness of the data collected, and participants’ rights, safety, and well-being. The type of PD will also be categorized according to these subtypes as recommended in Mohan et al 2016.^**12**^

1. Inclusion/exclusion criteria
2. Informed consent
3. Concomitant medication
4. Subject visit schedule
5. Study procedures/assessments
6. Treatment administration
7. SAE notification/safety procedure
8. Data protection and privacy
9. Other

Prior to datalock, all PDs were categorized as Major or Minor. Major PDs were defined as:

- Randomized though ineligible as study treatment has absolute contraindication
- Randomized though did not have a history of ICH
- Received the wrong treatment
- Concomitant medication used that had an absolute contraindication, or resulted in the total medication dose being above the maximal recommended dose
- No patient consent obtained
- Any other reasons that would jeopardise patient safety All other PDs were categorized as minor.

### 5.4 Analysis of the primary endpoint

#### 5.4.1 Primary analysis

The primary endpoint, time from randomization to first occurrence of recurrent stroke (whether ischemic, ICH or undifferentiated), will be summarized using Kaplan-Meier methods. The effect of the intervention will be estimated as the cause-specific HR and 95%CI obtained from a Cox proportional hazard model. Observations will be censored at the known time of death or when a participant was last known to be alive and free of event, thus estimating the risk of stroke in participants who are still alive.^15^ Stratification factors will be included as fixed covariates.

In case of clear violation of the proportional hazard assumption, as assessed visually with a plot of log(-log(survival)) versus log(time), differences in survival will be assessed using a log-rank test as well as by introducing a time-dependent variable to the Cox model.

As a sensitivity analysis, deaths due to any cause other than stroke will be treated as a competing risk.^**14**^ The analysis will be performed using a Fine and Gray competing risk model of the sub-distribution hazard, adjusted for stratification variables.^**13**^

#### 5.4.2 Adjusted analyses

Adjusted analyses of the primary endpoint will be performed by adding the following covariates to the Cox model: sex, pre-morbid level of function on the smRS, history of hypertension, history of cardiac disease, history of diabetes mellitus, level of education (<12 or ≥12 years). The adjusted treatment effect will be reported as HR and 95%CI.

#### 5.4.3 Subgroup analyses

Heterogeneity of treatment on the primary endpoint will be assessed in the pre-defined subgroups that are listed below. However, subgroups where there are fewer than 20% of participants will not be included in forest plot graphics given that the data are not informative due to the small sample size. Instead, these data will be included as footnotes to relevant figures and tables.

- Country of recruitment (Sri Lanka vs. other)
- Age (<60 vs ≥60 years)
- Sex (male vs female)
- Race – Asian, White, Black, Other
- Ethnicity – Hispanic or Latino, not Hispanic or Latino
- Hypertension status at screening – BP <140/90 vs BP ≥140/90 mmHg
- Background BP lowering intensity of treatment at screening (nil vs. 1 vs. ≥2 antihypertensive agents)
- Time since index event (<6 weeks vs ≥6 weeks)
- Presumed etiological (location) subtype of index ICH event (deep vs. cortical vs other)

The analysis for each subgroup will be performed by adding the subgroup variable as well as its interaction with the intervention as fixed effects to the main model (see Section 5.4.1). Within each subgroup, summary measures will include frequency, HR and standard error (SE) within each treatment arm, as well as the HR for treatment effect with a 95%CI. The results will be displayed on a forest plot (Figure 5) with the p-value for heterogeneity corresponding to the interaction term between the intervention and the subgroup variable.

Additional subgroup analyses will be performed as part of a sensitivity analysis to assess the comparability of IMP formulations (Section 5.8)

#### 5.4.4 Treatment of missing data

Given the nature of the primary endpoint (recurrent stroke), there is no reasonable way to assess whether some events are potentially missing (ie. unreported). We will therefore perform the analysis assuming that all events that occurred were reported, and without imputation of missing data. All reported events will be included in the time-to-first-event analysis with every participant censored at the known time of death, or the time when the participant was last known to be alive and free of an event. See Section 5.4.1 for more details about the primary analysis. The same approach will be used for every secondary survival endpoint.

### 5.5 Analyses of secondary endpoints

#### 5.5.1 Time to first occurrence of MACE

For time to first occurrence of MACE, we will use the same approach as the one used for the primary analysis (see Section 5.4.1). Participants not experiencing MACE events will be censored at the last date known to be alive and event free. No further adjustment or subgroup analysis will be performed.

#### 5.5.2 Time to CV mortality

For time to CV mortality, we will use the same approach as the one used for the primary analysis (see Section 5.4.1). Participants not experiencing CV death will be censored at the known time of death (if unrelated to CV events) or the last date known to be alive and event free. No further adjustment or subgroup analysis will be performed.

#### 5.5.3 Hypertension control at 6 months

Hypertension control will be defined as a SBP measurement <130 mmHg. Hypertension control will be summarized using descriptive statistics at each measured visit.

The primary analysis of hypertension control will consist in comparing the occurrence of hypertension control at 6 months of follow-up between treatment groups using logistic regression, including stratification factors (i.e. country of recruitment, age (<65 vs. ≥65 years) and SBP at randomization as a continuous variable) as fixed effects. Participants missing the 6-month follow-up assessment will be considered as not having controlled hypertension for the purpose of the primary analysis.

### 5.6 Safety outcomes

Serious adverse events (SAEs) and adverse events of special interest (AESIs) will be grouped by type and category (using MedRA system organ class and preferred terms for SAEs). The total number of events as well as the number and proportion of participants experiencing at least one event will be tabulated.

### 5.7 Analysis of other outcomes

No formal significance tests will be applied to these outcomes. Instead, the focus will be on description and estimation of overall differences between the two randomized arms using all available data. We understand that the amount of data available will vary between participants depending on when they entered the study and when they were last followed up (censoring). Assuming that reasons for censoring are reasonably well-balanced between the two randomized arms, these analyses based on all data available should be unbiased.

#### 5.7.1 Time to event endpoints

Other time to event endpoints will be analyzed in a similar manner to the primary endpoint using Kaplan-Meier plots and Cox proportional hazards models, adjusted for stratification factors as covariates. Point estimates for HRs and 95%CI will be presented.

For the following time to event endpoints, observations will be censored at the known time of death unrelated to the specified endpoint or, for those last known to be alive, at the last time a participant was known to be event free:

- Time to ICH
- Time to ischemic stroke
- Time to fatal stroke
- Time to stroke of undifferentiated origin
- Time to components of the MACE outcome (to be analyzed separately)

Time to all-cause mortality: Participants will be censored on the date last known to be alive.

#### 5.7.2 Win ratio analysis

Primary and secondary endpoints will be analyzed jointly using a win ratio approach which ranks all possible outcomes in order of importance. The following hierarchy will be used:

1. CV death
2. Non-CV death
3. Nonfatal stroke
4. Dementia
5. Non-fatal MI
6. Dependency, defined by smRS 3 to 5
7. MCI

Every participant randomized to the intervention arm will be paired with every participant randomized to the control arm. For illustrative purposes, if we had 130 patients randomized to one arm and 127 to the other, we would compare a total of 16,510 pairs (130 × 127). Within each pair, we will compare outcomes in a hierarchical fashion until a winner has been determined. If no winner can be declared after comparing all outcomes, the pair will be tied. We will start by comparing CV deaths to determine a “winner”. The winner will be the participant with the longest time to CV death. If neither participant died of a CV cause and a winner cannot be declared on CV death alone, we will then proceed to the next step and compare time to non-CV death. Again, the winner will be the participant with the longest time to non-CV death. Only events that occur during the time that both participants are in the study will be used (e.g. if one participant withdraws after 1 year, only the events occurring in the first year will be used to compare the pair). The approach will proceed in a stepwise fashion, moving to the next outcome in the hierarchy, until a decision has been reached for every pair. The full decision process is described in the table below. The win ratio will then be calculated as the proportion of “intervention group winners” (the participants in the intervention group who had a better outcome) divided by the proportion of “control winners” (the participants in the control group who had a better outcome).

To account for the stratification factors used in the randomization, the win ratio approach will be stratified by country, age and baseline BP, following the method proposed by Dong et al.^15^ This means that patients from the Triple Pill arm from one stratum will only be paired (and compared) to patients from the placebo from the same stratum (i.e. same country, same age group and same baseline BP category). To complete the interpretation, we will also compute the stratified win odds and net benefit^.16,17^ 95%CIs will be computed using the delta method as described by Dong et al.^17^ Calculations will be performed using the R package WINS developed by Cui and Huang.^18^

To account for the fact that patients might have different follow-up times, comparisons will only be made based on the shortest follow-up time in the pair. For example, if one patient from the pair was followed for 12 months and the other for 18 months, we will only determine the winner using data collected in the first 12 months post-randomization. For time-to-event outcomes (e.g. time to CV death), this means that the analysis will be censored at the shortest follow-up time. For other outcomes (dementia, dependency and MCI), the comparison will be performed at the latest common visit. In case of missing data from one or two subjects at the latest common visit, the pair will be declared a tie (unable to declare a winner).

| Step | Outcome | Decision rule |
| --- | --- | --- |
| 1 | Time to CV death | The winner is the patient with the longest time to CV death. If neither died of a CV cause, go to step 2. If both died of a CV cause on the same day, go to step 3. |
| 2 | Time to non-CV death | The winner is the patient with the longest time to non-CV death. If neither died of a non-CV cause or if both died of a non-CV cause on the same day, go to step 3. |
| 3 | Time to stroke | The winner is the patient with the longest time to first stroke. If neither had a stroke or if both patients had their first stroke on the same day, go to step 4. |
| 4 | Dementia | The winner is the patient with no dementia at the latest common visit. If neither or both patients had dementia, go to step 5. |
| 5 | Time to MI | The winner is the patient with the longest time to first MI. If neither had a MI or if both patients had their first MI on the same day, go to step 6. |
| 6 | Dependency | The winner is the patient with no dependency defined by mRS 3 or more at the latest common visit. If neither or both patients had dependency, go to step 7. |
| 7 | MCI | The winner is the patient with no MCI at the latest common visit. If neither or both patients had MCI, the pair is declared a tie. |

#### 5.7.3 Dementia-free survival

Dementia-free survival will be defined as being alive and without a dementia diagnosis at the time of the last visit. Summary statistics of the number assessed and number and percentage with dementia at the end of follow-up will be reported.

Dementia-free survival will be analyzed using a binomial regression with an offset term to account for variable follow-up. All available information will be included in the regression model for participants with recorded deaths or a non-missing dementia assessment. Fixed effects will include the randomized treatment as well as the stratification variables of country, age at recruitment (<65 vs ≥65 years), and baseline SBP measurements (<140 vs ≥140 mmHg). We will thus derive a yearly incidence rate of death or dementia which will be compared between treatment arms using an incidence rate ratio and 95%CI. No further adjustment or subgroup analysis will be applied.

Sensitivity analyses will include:

- Binomial regression model as specified for the primary analysis of dementia-free survival including education level (secondary or above vs other) as a fixed effect;
- A binomial regression model as specified for the primary analysis, but imputing all missing observations as dementia cases;
- A tipping point analysis as a sensitivity analysis and to assess the robustness of the findings based on complete cases.

#### 5.7.4 MCI

MCI will be defined as being free of MCI and alive at the time of the last visit. Summary statistics of the number assessed and the number and percentage with MCI at the end of follow-up will be reported. In addition, MCI will be analyzed using the same approach as dementia-free survival, using a binomial regression with an offset term to account for variable follow-up (see Section 5.7.3). All available information will be included in the regression model for participants with non-missing MCI assessments. Fixed effects will include the randomized treatment as well as the stratification variables of country, age at recruitment (<65 vs ≥65 years) and baseline SBP measurements (<140 vs ≥140 mmHg). As a sensitivity analysis, missing MCI assessments will be imputed as MCI cases.

#### 5.7.5 Composite of dementia and MCI

The composite of dementia and MCI will be defined as being free of MCI or dementia and alive at the time of the last visit. The composite will be summarized as the number assessed and the number and percentage with dementia or MCI at the end of follow-up. In addition, the composite will be analyzed using the same approach as dementia-free survival, using a binomial regression with an offset to account for variable follow-up (see Section 5.7.3). All available information will be included in the regression model for participants with non-missing dementia and/or MCI assessments. Fixed effects will include the randomized treatment as well as the stratification variables of country, age at recruitment (<65 vs ≥65 years) and baseline SBP (<140 vs ≥140 mmHg). As a sensitivity analysis, missing MCI or dementia assessments will be imputed as cases.

#### 5.7.6 Cognitive decline – MoCA, BMET

For the analysis of MoCA, a total score will be calculated by aggregating scores from the individual items in the questionnaire and summarized using descriptive statistics at each visit with assessments. Assuming a normal distribution and a link identity function, a repeated-measure linear mixed model will be used to estimate the effect of the intervention overall and by visit. The model will include all post-randomization values after reallocation of the EOS visit (see Section 4.6). Fixed effects will include treatment, visit, treatment by visit interaction term as well as the stratification variables of country, age at recruitment (<60 vs ≥60 years) and baseline SBP (<140 vs ≥140 mmHg) as well as the MoCA score at randomization. Within-patient correlations will be modelled using a repeated patient effect assuming a compound-symmetry (exchangeable) structure. The overall effect of the intervention will be estimated as the mean difference and 95%CI between the intervention and control arm over the entire follow-up (i.e. by combining data obtained across all visits). The effect of the intervention at specific time points (e.g. at month 12) will be estimated from the same model using contrasts. A similar analysis will be applied to the analysis of BMET in the subset of participants in which BMET was collected. For both MoCA and BMET, no missing data imputation, further adjustment or subgroup analysis are planned.

#### 5.7.7 Survival free of severe disability (mRS 0-2)

A binary analysis of the mRS will be performed by dichotomising the mRS as either ‘dead or dependent’ (scores 3-6) or ‘independent’ (scores 0-2) and summarized at each visit with mRS assessments using descriptive statistics.

The primary analysis of disability-free survival at the end of follow-up will be analyzed using all reported information. A binomial regression where an “event” is defined as death or dependent mRS score (scores 3-6) at the end of follow-up visit. If the end of follow-up is missing and the participant is known to be alive, the last recorded mRS observation score will be used. The variable follow-up time will be accounted for using the log(follow-up) as an “offset”. Fixed effects will include the randomized treatment as well as the stratification variables of country, age at recruitment (<65 vs ≥65 years) and baseline SBP measurements (<140 vs ≥140 mmHg). We will thus derive an incidence rate ratio and 95%CI to compare treatment groups.

#### 5.7.8 Physical functional assessment using the smRS (Ordinal analysis)

Physical function assessment using the smRS summarized using descriptive statistics at each visit.

The distributions of scores on the smRS between randomized groups will be compared using repeated-measure ordinal logistic regression with a cumulative logit link function. The model will include all post-randomization smRS values after reallocation of the EOS visit (see Section 4.6). Fixed effects will include treatment, visit, treatment by visit interaction term as well as the stratification variables of country, age at recruitment (<60 vs ≥60 years) and baseline SBP measurements (<140 vs ≥140 mmHg). Within-patient correlations will be modelling using a repeated patient effect assuming a compound-symmetry (exchangeable) structure. The overall effect of the intervention will be estimated as the OR and 95%CI between the intervention and control arm over the entire follow-up, i.e. by combining data obtained across all visits. The effect of the intervention at specific time points (e.g. at month 12) will be estimated from the same model using contrasts.

We will test the proportional-odds assumption using a score test. In case of violation, we will still proceed with the analysis and interpret the intervention OR as an average effect across all smRS levels but with the understanding that it may not be constant across all levels. This will be complemented by a graphical assessment of shifts across categories using bar plots as well as a binary analysis.

#### 5.7.9 Health related quality of life – EQ-5D-3L

For the analysis of EQ-5D, a health utility score will be calculated for each participant at each visit using EuroQol EQ-5D-3L calculation specifications and summarized using descriptive statistics. The health utility score will be analyzed using a repeated-measure linear mixed model (i.e. normal distribution and identity link). The model will include all post-randomization utility scores after reallocation of the EOS visit (see Section 4.6). Fixed effects will include treatment, visit, treatment by visit interaction term as well as the stratification variables of country, age at recruitment (<60 vs ≥60 years) and SBP at randomization (<140 vs ≥140 mmHg) and the health utility score at randomization as a continuous covariate. Within-patient correlations will be modelled using a repeated patient effect assuming a compound-symmetry (exchangeable) structure. The overall effect of the intervention will be estimated as the mean difference and 95%CI between the intervention and control arm over the entire follow-up (i.e. by combining data obtained across all visits). The effect of the intervention at specific time points (e.g. at month 12) will be estimated from the same model using contrasts For each of the 5 dimensions, we will only perform descriptive analyses, reporting the number and proportion of participants in each category using tabular and graphical summaries. No missing data imputation, further adjustment or subgroup analysis are planned.

#### 5.7.10 BP

The actual values and change in BP from randomization to post-randomization will be summarized using descriptive statistics at each visit.

Hypertension control throughout follow-up is defined as the mean of all post-randomization SBP measures being <130 mmHg. The number and percentage of participants achieving control throughout follow-up will be summarized.

Hypertension control at the end of follow-up, defined as SBP <130mmHg at the final follow-up visit, will be analyzed using all available information. No imputation will be applied. A binomial regression where “success” is defined as hypertension control at the last recorded visit will be performed. The variable follow-up time will be accounted for using the log(follow-up) as an “offset”. Fixed effects will include the randomized treatment as well as the stratification variables of country, age at recruitment (<65 vs ≥65 years) and baseline SBP measurements (<140 vs ≥140 mmHg). We will thus derive an incidence rate ratio and 95%CI to compare treatment groups.

Change in SBP and DBP will be analyzed separately using repeated-measure linear mixed models (i.e. normal distribution and identity link). The model will include all post-randomization BP values after reallocation of the EOS visit (see Section 4.6). Fixed effects will include treatment, visit, treatment by visit interaction term as well as the stratification variables of country, age at recruitment (<60 vs ≥60 years) and SBP at randomization as a continuous variable. Within-patient correlations will be modelling using a repeated patient effect assuming a compound-symmetry (exchangeable) structure. The overall effect of the intervention will be estimated as the mean difference and 95%CI between the intervention and control arm over the entire follow-up, i.e. by combining data obtained across all visits. The effect of the intervention at specific time points (e.g. at Month 12) will be estimated from the same model using contrasts.

Time at target during follow-up is defined as the estimated proportion of follow-up time for each participant at SBP <130mmHg, based on available post-randomization SBP measures and according to methods previously described. Time at target will be computed using linear interpolation, as described by Chung et al (2018).^19^ The average proportion of time at target will be reported by treatment arm and between-arm difference will be estimated using a linear model (normal distribution and identity link). Fixed effects will include the randomized treatment as well as the stratification variables of country, age at recruitment (<60 vs ≥60 years) and SBP at randomization as a continuous variable. The effect of the intervention will be estimated as the mean difference and 95%CI.

Given that there is expected to be much greater power for mean SBP difference endpoint and the essential role of BP reduction in stroke prevention, subgroup analyses, as described in Section 5.4.3 will be conducted for the SBP difference, by finer subgroup categories (e.g., for each country separately, pooling together those randomizing <50 patients; for <50, 50-59, 60-69 and 70+ years of age).

### 5.8 Sensitivity analyses according to IMP

Until 6 June 2022, randomized participants received IMP comprised of over-encapsulation of existing on market monotherapies. After this date, GMRx2-dose triple 1/2 tablet or matching placebo was used. Overall, 314 randomized participants received triple pill/matching placebo only, 641 received triple pill/placebo with a switch to GMRx2/placebo, and 715 received GMRx2/matching placebo only.

To assess any impact of the IMP, subgroup analyses of the primary endpoint will be performed. The subgroup will be defined according to the study medication formulation allocated at the time of randomization (i.e. over-encapsulated [some of whom switched over to GMRx2 during follow-up] vs GMRx2). The approach is similar to other subgroup analyses (see Section 5.4.3) where the regression model will be repeated including the fixed effect for the formulation period (i.e., triple pill/placebo versus GMRx2/placebo) and an interaction between the formulation period and treatment assignment. The interaction term will inform whether the treatment effect observed differs by formulation period.

A stratified analysis of BP endpoints will be performed according to the same subgroups. This will be done by rerunning the analyses of BP change and BP control described above separately for the two IMP subgroups (i.e. for those who started with the over-encapsulated version vs for those who started on GMRx2). The same analysis methods will be performed assessing change in BP from baseline to week 4 of follow-up for the two pre-specified formulation period subgroups.

Finally, a descriptive analysis will be conducted of the between group BP differences at the clinic visit at which trial medication was switched from triple pill to GMRx2, which was the last visit at which any BP differences were due to triple pill efficacy. This will be presented alongside the between group BP difference at the next clinic visit, which is the first visit at which any BP differences were due to GMRx2 efficacy. Descriptive data will also be presented on temporal trends of BP differences, and any differential follow-up between groups at each clinic visit.

### 5.9 MRI substudy

The aim of this substudy is to determine the effectiveness of Triple Pill vs. placebo on markers of cerebral small vessel disease (CSVD) – white matter hyperintensities (WMH), cerebral microbleeds (CMB), lacunes, perivascular spaces, cerebral atrophy, and cortical superficial siderosis (CSS) – in patients with ICH.

Centers capable of acquiring routine clinical MRI flair sequences, ideally with 3D but for pragmatic reasons 2D scans are also allowed for identification, will be used where there is also interest and capacity, and ability to undertake high-quality MRI scanning with a reasonable volume of patients. Coordinating personnel will collect and send the necessary MRI data to the Magnetic Resonance-Coordinating Centre (MR-CC), based at the Sydney Neuroimaging Analysis Centre (SNAC) in the Brain and Mind Centre (BMC) of the University of Sydney, Australia. The data are sent electronically.

Patients identified as being eligible for MRI will have this test performed at the penultimate or final study visit time point. It is anticipated that approximately 60% of patients will be eligible for MRI to achieve a sample size of 750 patients. All data collected will be analysed centrally. The list of metrics to be produced according to the type of scan are outlined below.

| Endpoint | Outcome | MRI image |  |  |  |
| --- | --- | --- | --- | --- | --- |
|  |  | T1 | Flair | SWI | DWI |
| Primary | Proportion of WML |  | X |  |  |
| Secondary | Recent small subcortical infarct |  |  |  | X |
|  | WMH |  | X |  |  |
|  | Lacune of presumed vascular origin |  | X |  |  |
|  | Cerebral microbleeds (CMB) |  |  | X |  |
|  | Cortical superficial siderosis (CSS) |  |  | X |  |

Most of the analyses will be done through an automated AI-based tool, iQ-solutions.^20,21^ Brain volumetrics will also be undertaken: whole brain volume, cortical gray matter and lobar volumes, using iQ-COG.

The primary analysis of WMH volume will be analyzed using all available information. Because of the skewed distribution for WMH volume, we will first apply an inverse hyperbolic sine transformation (asinh), which is similar to a log transformation but can accommodate values of zero. Linear mixed models, including random effects for participant, MRI facility, country, intracranial volume, age at recruitment (<65 vs ≥65 years), and baseline SBP measurements (<140 vs ≥140 mmHg), will be used to estimate the difference in WMH volume between the treatment groups. The variable follow-up time will be accounted for using the log(follow-up) as an “offset”. Fixed effects will include the randomized treatment. The overall effect of the intervention will be estimated as the mean difference and 95%CI between the intervention and control arm. A sensitivity analysis will include additional covariates of brain volume

Exploratory analysis will be undertaken for the other MRI markers of CSVD and their relation to cognitive impairment and physical function.

## Data Availability

All data produced in the present study are available upon reasonable request to the Principal Investigator, Professor Craig Anderson

## 7 Revision History: Summary of Changes

The following changes were implemented in Version 2.0.

| Section | Description of Change |
| --- | --- |
| General | Language updates are executed throughout the document to align terminology and spelling. |
| 1 | Added Version 2.0 to Revision history, reordered the contributors and authors of this analysis plan according to relative contributions with the most recent modifications.<br>Identified that changes were made to the SAP version 2.0 that no longer align with the protocol-specified analyses. Modifications were made to clarify the endpoints and conclusions, the analyses to be performed, and to align with FDA comments received May 2025. |
| 2.2 | Study population section was made more general to avoid confusion given the later-described changes to the sample size. |
| 2.4 | Primary study outcome is unchanged.<br>Secondary and exploratory study outcomes were reordered to clearly identify the importance and conclusions to be drawn. Secondary efficacy outcomes are identified and align with the planned statistical hypothesis testing and corresponding type I error control. The remaining outcomes are identified as exploratory. |
| 4.1 | The sample size section was clarified to identify the modification from the planned sample size was due to challenges in recruitment as well as observed results from a prior, published study (see Section for detail). It is also clarified that the change in sample size was determined in a blinded fashion, without access to unblinded data. |
| 4.3 | The role of the independent Data Safety Monitoring Board is clarified. Access to unblinded information and execution of the unblinded interim analysis was performed by designated unblinded personnel. The study team remains blinded. |
| 4.4 | The multiplicity adjustment section is modified to align with the reordered and reduced number of secondary efficacy endpoints. |
| 4.7 | The missing data handling section was added to the SAP version 2.0. |
| 5.1 | An analysis of the counts and reasons for discontinuation after the single blind active run-in is added to the analysis. Additional clarifications to analysis methods are added, including the handling of missing data and censored observations. |
| 5.4 | Sub-sections within 5.4 are reorganized to align with the reordered efficacy endpoints. |
| 5.4.1 | The primary analysis method of the primary endpoint is modified to use Kaplan-Meier methods and a Cox Proportional Hazards model. A sensitivity analysis using the Fine and Gray competing risk analysis is planned. |
| 5.5 | Analysis methods for the three secondary efficacy endpoints are described. |
| 5.7 | Analysis of other exploratory endpoints are detailed and are listed in the order of the efficacy outcomes. |
| 5.7.2 | The win ratio analysis method is described, including the planned comparison of all participants within stratum. To account for different follow-up times, comparisons between subjects will be executed at the earlier (shorter) of the two follow-up periods, as specified within the section. |
| 5.7.3 | Clarifications are made to the dementia-free survival analysis section. |
| 5.7.4 | Clarifications are made to the MCI analysis section. |
| 5.7.5 | Clarifications are made to the dementia and MCI composite analysis section. |
| 5.7.7 | The analysis of survival free of severe disability is described. |
| 5.7.10 | Blood pressure summaries and analysis are described, including descriptive summaries by visit, the analysis of exploratory endpoints, and planned subgroup analyses planned. |
| 5.8 | A descriptive analysis is added to summarize differences between observed blood pressure measurements in clinic when subjects were treated with the triple pill versus GMRx2. |

The following changes were implemented in Version 3.0.

| Section | Description of Change |
| --- | --- |
| 5.3 | Inclusion of Protocol Deviation definitions |
| 5.9 | Inclusion of analysis of MRI data |
| 8 | Inclusion of listing of planned publications |
| 9 | Inclusion of figure and table shells |

## 8 Proposed publications

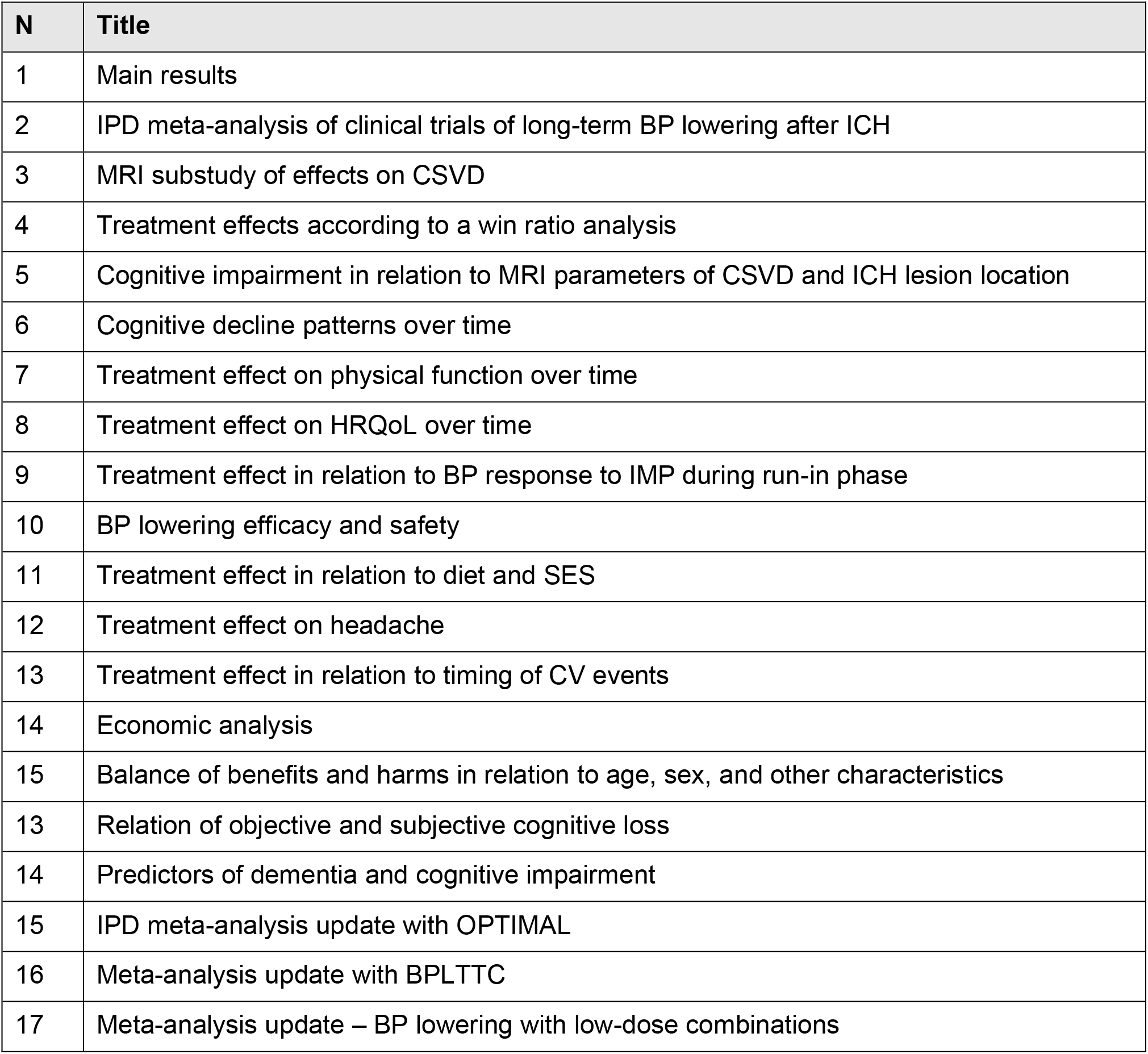

| N | Title |
| --- | --- |
| 1 | Main results |
| 2 | IPD meta-analysis of clinical trials of long-term BP lowering after ICH |
| 3 | MRI substudy of effects on CSVD |
| 4 | Treatment effects according to a win ratio analysis |
| 5 | Cognitive impairment in relation to MRI parameters of CSVD and ICH lesion location |
| 6 | Cognitive decline patterns over time |
| 7 | Treatment effect on physical function over time |
| 8 | Treatment effect on HRQoL over time |
| 9 | Treatment effect in relation to BP response to IMP during run-in phase |
| 10 | BP lowering efficacy and safety |
| 11 | Treatment effect in relation to diet and SES |
| 12 | Treatment effect on headache |
| 13 | Treatment effect in relation to timing of CV events |
| 14 | Economic analysis |
| 15 | Balance of benefits and harms in relation to age, sex, and other characteristics |
| 13 | Relation of objective and subjective cognitive loss |
| 14 | Predictors of dementia and cognitive impairment |
| 15 | IPD meta-analysis update with OPTIMAL |
| 16 | Meta-analysis update with BPLTTC |
| 17 | Meta-analysis update – BP lowering with low-dose combinations |

## Notes

### Competing Interest Statement

AR is seconded part-time to work for George Medicines (GM), which is partly owned by George Health Enterprises (GHE), the social enterprise arm of The George Institute for Global Health (TGI), which holds patents for low-dose fixed-dose combination products for the treatment of hypertension and diabetes; AR is listed as one of the inventors (US 10,369,15; US 10,799,487; US 10,322,117; US 11,033,544). GHE and GM have received funding from public and private investors to conduct the research required for regulatory approval of cardiovascular combination products; AR does not have a financial interest in these patent applications or investments. CA is a consultant for Auzone BioTech China. RB is employed by Veristat. LB and SS report no conflicts of interest.

### Clinical Trial

NCT02699645

### Funding Statement

This study was funded by The National Health and Medical Research Council (NHMRC) of Australia (Grants 1149987 and 1103886) and PROADI SUS - Ministry of Health of Brazil and Hospital Moinhos de Vento (NUP number 25000.209767/2018-61).

### Author Declarations

The Protocol X16-0082 & 2019/ETH06459 (GI-AU-NMH-2016-01, Version 6.0, Date 6 December 2022): Triple therapy prevention of Recurrent Intracerebral Disease EveNts Trial was approved by the Research Ethics and Governance Office of Sydney Local Area Health District, office at the Royal Prince Alfred Hospital, CAMPERDOWN NSW Australia 2050

