## Supplemental files for "Triple therapy prevention of Recurrent Intracerebral Disease EveNts Trial (TRIDENT)"

Docusign Envelope ID: CD9CCBFF-A0AC-4C1B-A386-8B23A4692D4E

**An investigator initiated and conducted, multicenter, international, double-blind, placebo-controlled, parallel-group, randomized controlled trial to determine the effectiveness of more intensive blood pressure control provided by a fixed low-dose of blood pressure lowering agents as a single pill combination 'Triple Pill' strategy on top of standard of care, on the time to first occurrence of recurrent stroke in patients with a history of stroke due to intracerebral hemorrhage.**

**Statistical Analysis Plan**

***Shells for tables and figures***

Version: 3.0

Date: 22 August 2025

**Authors:**

Laurent Billot, The George Institute, UNSW Sydney, Australia

Sana Shan, The George Institute, UNSW Sydney, Australia

Anthony Rodgers, The George Institute, UNSW Sydney, Australia

Craig Anderson, The George Institute, UNSW Sydney, Australia

Robin Bliss, Veristat, Southborough, Massachusetts, USA

Docusign Envelope ID: CD9CCBFF-A0AC-4C1B-A386-8B23A4692D4E

| Name | Signature | Date |
|---|---|---|
| Prof Laurent Billot | Signed by:<br>Signer Name: Laurent Billot<br>Signing Reason: I approve this document<br>Signing Time: 24 August 2025 \| 1:51:16 PM PDT<br>F638154CB59E4B2E9E2318317ED037EC | 24 August 2025 |
| Ms Sana Shan | Signed by:<br>Signer Name: Sana Shan<br>Signing Reason: I am the author of this document<br>Signing Time: 24 August 2025 \| 2:48:49 PM PDT<br>B0DC73DEF86D49B79B0C5BFE31A27106 | 24 August 2025 |
| Prof Anthony Rodgers | Signed by:<br>Signer Name: Anthony Rodgers<br>Signing Reason: I am the author of this document<br>Signing Time: 24 August 2025 \| 3:56:33 AM PDT<br>5742BF25E84B47E0B7F2F5005A5644F8 | 24 August 2025 |
| Prof Craig Anderson | Signed by:<br>Signer Name: Craig Anderson<br>Signing Reason: I am the author of this document<br>Signing Time: 25 August 2025 \| 5:21:53 AM AEST<br>63946B7DA1724B929EC1030118682790 | 25 August 2025 |
| Dr Robin Bliss | Signed by:<br>Signer Name: Robin Bliss<br>Signing Reason: I approve this document<br>Signing Time: 24 August 2025 \| 6:35:24 AM EDT<br>CE65E78D368F4090866A1C16992D4794 | 24 August 2025 |

Docusign Envelope ID: CD9CCBFF-A0AC-4C1B-A386-8B23A4692D4E

### Figure and Table shells

#### Figure 1: Consort flowchart

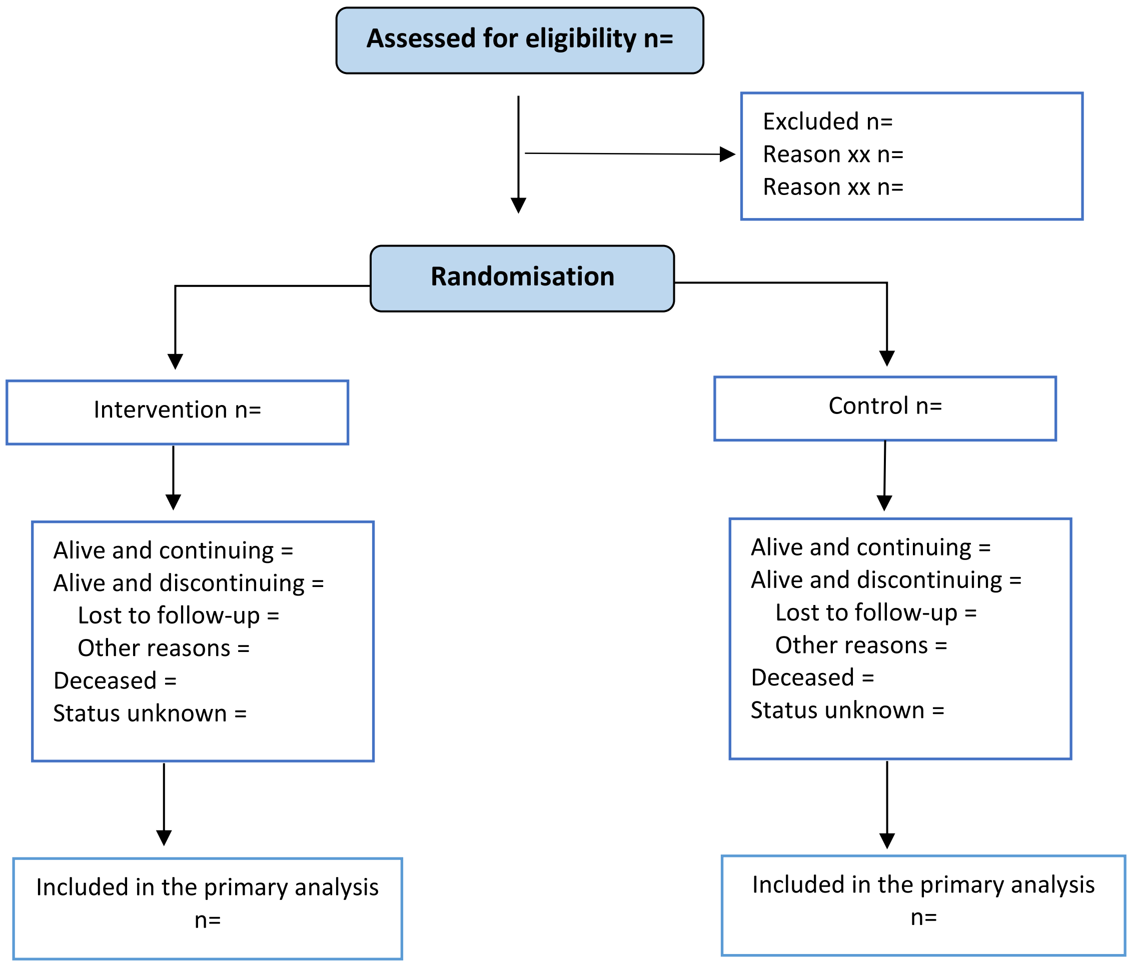

Docusign Envelope ID: CD9CCBFF-A0AC-4C1B-A386-8B23A4692D4E

**Table 1. Enrolment by country and site**

| Country | Number of subjects | |
|---|---|---|
| Centre | Screened N=xx | Randomised N=xx |
| Australia | xx(xx.x%) | xx(xx.x%) |
| Centre 1 | xx(xx.x%) | xx(xx.x%) |
| Centre 2 | xx(xx.x%) | xx(xx.x%) |
| etc… | xx(xx.x%) | xx(xx.x%) |
| England | xx(xx.x%) | xx(xx.x%) |
| Centre 1 | xx(xx.x%) | xx(xx.x%) |
| Centre 2 | xx(xx.x%) | xx(xx.x%) |
| etc… | xx(xx.x%) | xx(xx.x%) |
| Netherlands | | |
| Centre 1 | xx(xx.x%) | xx(xx.x%) |
| Centre 2 | xx(xx.x%) | xx(xx.x%) |
| etc… | xx(xx.x%) | xx(xx.x%) |
| Sri Lanka | xx(xx.x%) | xx(xx.x%) |
| Centre 1 | xx(xx.x%) | xx(xx.x%) |
| Centre 2 | xx(xx.x%) | xx(xx.x%) |
| etc… | xx(xx.x%) | xx(xx.x%) |
| Georgia | xx(xx.x%) | xx(xx.x%) |
| Centre 1 | xx(xx.x%) | xx(xx.x%) |
| Centre 2 | xx(xx.x%) | xx(xx.x%) |
| etc… | xx(xx.x%) | xx(xx.x%) |
| Brazil | xx(xx.x%) | xx(xx.x%) |
| Centre 1 | xx(xx.x%) | xx(xx.x%) |
| Centre 2 | xx(xx.x%) | xx(xx.x%) |
| etc… | xx(xx.x%) | xx(xx.x%) |
| Nigeria | xx(xx.x%) | xx(xx.x%) |
| Centre 1 | xx(xx.x%) | xx(xx.x%) |
| Centre 2 | xx(xx.x%) | xx(xx.x%) |
| etc… | xx(xx.x%) | xx(xx.x%) |

Docusign Envelope ID: CD9CCBFF-A0AC-4C1B-A386-8B23A4692D4E

**Table 2. Patient disposition**

| Status | Intervention | Control | Total |
|---|---|---|---|
| **Eligible for randomization** | N=xxx | N=xxx | N=xxx |
| Intolerance of run-In/rerun-in medication | xx (xx.x%) | xx (xx.x%) | xx (xx.x%) |
| **Randomized** | N=xxx | N=xxx | N=xxx |
| **Overall numbers** | | | |
| Alive and completed the study | xx (xx.x%) | xx (xx.x%) | xx (xx.x%) |
| Discontinued | xx (xx.x%) | xx (xx.x%) | xx (xx.x%) |
| Death | xx (xx.x%) | xx (xx.x%) | xx (xx.x%) |
| Lost to follow-up | xx (xx.x%) | xx (xx.x%) | xx (xx.x%) |
| Withdrew consent | xx (xx.x%) | xx (xx.x%) | xx (xx.x%) |
| Investigator's decision | xx (xx.x%) | xx (xx.x%) | xx (xx.x%) |
| Other | xx (xx.x%) | xx (xx.x%) | xx (xx.x%) |

Docusign Envelope ID: CD9CCBFF-A0AC-4C1B-A386-8B23A4692D4E

**Table 3. Follow-up mode of assessment**

| Timepoints | Intervention | Control | Total |
|---|---|---|---|
| **6 Weeks** | **N=xxx** | **N=xxx** | **N=xxx** |
| Clinic | xx (xx.x%) | xx (xx.x%) | xx (xx.x%) |
| Phone call | xx (xx.x%) | xx (xx.x%) | xx (xx.x%) |
| Phone call with other | xx (xx.x%) | xx (xx.x%) | xx (xx.x%) |
| Information obtained via medical records | xx (xx.x%) | xx (xx.x%) | xx (xx.x%) |
| Not done | xx (xx.x%) | xx (xx.x%) | xx (xx.x%) |
| **6 Months** | **N=xxx** | **N=xxx** | **N=xxx** |
| Clinic | xx (xx.x%) | xx (xx.x%) | xx (xx.x%) |
| Phone call | xx (xx.x%) | xx (xx.x%) | xx (xx.x%) |
| Phone call with other | xx (xx.x%) | xx (xx.x%) | xx (xx.x%) |
| Information obtained via medical records | xx (xx.x%) | xx (xx.x%) | xx (xx.x%) |
| Not done | xx (xx.x%) | xx (xx.x%) | xx (xx.x%) |
| **12 Months** | **N=xxx** | **N=xxx** | **N=xxx** |
| Clinic | xx (xx.x%) | xx (xx.x%) | xx (xx.x%) |
| Phone call | xx (xx.x%) | xx (xx.x%) | xx (xx.x%) |
| Phone call with other | xx (xx.x%) | xx (xx.x%) | xx (xx.x%) |
| Information obtained via medical records | xx (xx.x%) | xx (xx.x%) | xx (xx.x%) |
| Not done | xx (xx.x%) | xx (xx.x%) | xx (xx.x%) |
| **18 Months** | **N=xxx** | **N=xxx** | **N=xxx** |
| Clinic | xx (xx.x%) | xx (xx.x%) | xx (xx.x%) |
| Phone call | xx (xx.x%) | xx (xx.x%) | xx (xx.x%) |
| Phone call with other | xx (xx.x%) | xx (xx.x%) | xx (xx.x%) |
| Information obtained via medical records | xx (xx.x%) | xx (xx.x%) | xx (xx.x%) |
| Not done | xx (xx.x%) | xx (xx.x%) | xx (xx.x%) |
| **24 Months** | **N=xxx** | **N=xxx** | **N=xxx** |
| Clinic | xx (xx.x%) | xx (xx.x%) | xx (xx.x%) |
| Phone call | xx (xx.x%) | xx (xx.x%) | xx (xx.x%) |
| Phone call with other | xx (xx.x%) | xx (xx.x%) | xx (xx.x%) |
| Information obtained via medical records | xx (xx.x%) | xx (xx.x%) | xx (xx.x%) |
| Not done | xx (xx.x%) | xx (xx.x%) | xx (xx.x%) |
| **Repeated for all follow-up measurements** | | | |

**Table 4. Baseline characteristics**

| Characteristics | Intervention | Control | Total |
|---|---|---|---|
| **Age (years)** | | | |
| N Mean (SD) | N xx.x (xx.x) | N xx.x (xx.x) | N xx.x (xx.x) |
| Median [Q1 – Q3] | xx.x [xx.x - xx.x] | xx.x [xx.x - xx.x] | xx.x [xx.x - xx.x] |
| **Age categories** | | | |
| <65 years | xx (xx.x%) | xx (xx.x%) | xx (xx.x%) |
| ≥65 years | xx (xx.x%) | xx (xx.x%) | xx (xx.x%) |
| **Sex** | N=xxx | N=xxx | N=xxx |
| Male | xx (xx.x%) | xx (xx.x%) | xx (xx.x%) |
| Female | xx (xx.x%) | xx (xx.x%) | xx (xx.x%) |
| **Weight (kg)** | | | |
| N_ Mean(SD) | N xx.x (xx.x) | N xx.x (xx.x) | N xx.x (xx.x) |
| Median [Q1 – Q3] | xx.x [xx.x - xx.x] | xx.x [xx.x - xx.x] | xx.x [xx.x - xx.x] |
| **BMI (kg/m$^2$)** | | | |
| N Mean(SD) | N xx.x (xx.x) | N xx.x (xx.x) | N xx.x (xx.x) |
| Median [Q1 – Q3] | xx.x [xx.x - xx.x] | xx.x [xx.x - xx.x] | xx.x [xx.x - xx.x] |
| **Baseline SBP categories** | | | |
| <140mmHg | xx (xx.x%) | xx (xx.x%) | xx (xx.x%) |
| ≥ 140mmHg | xx (xx.x%) | xx (xx.x%) | xx (xx.x%) |
| BP monitoring at home | xx (xx.x%) | xx (xx.x%) | xx (xx.x%) |
| **Used regularly (at least 1/month)** | xx (xx.x%) | xx (xx.x%) | xx (xx.x%) |
| **Ethnicity**(1) | | | |
| Caucasian | xx (xx.x%) | xx (xx.x%) | xx (xx.x%) |
| African descent | xx (xx.x%) | xx (xx.x%) | xx (xx.x%) |

| | | | |
|---|---|---|---|
| Arabic | xx (xx.x%) | xx (xx.x%) | xx (xx.x%) |
| Hispanic / Latino | xx (xx.x%) | xx (xx.x%) | xx (xx.x%) |
| East Asian | xx (xx.x%) | xx (xx.x%) | xx (xx.x%) |
| South Asian | xx (xx.x%) | xx (xx.x%) | xx (xx.x%) |
| South-East Asian | xx (xx.x%) | xx (xx.x%) | xx (xx.x%) |
| Australian Aboriginal | xx (xx.x%) | xx (xx.x%) | xx (xx.x%) |
| Maori | xx (xx.x%) | xx (xx.x%) | xx (xx.x%) |
| Middle Eastern descent | xx (xx.x%) | xx (xx.x%) | xx (xx.x%) |
| Torres Strait Islander | xx (xx.x%) | xx (xx.x%) | xx (xx.x%) |
| Pacific Islander | xx (xx.x%) | xx (xx.x%) | xx (xx.x%) |
| Indigenous Brazilian | xx (xx.x%) | xx (xx.x%) | xx (xx.x%) |
| Israeli | xx (xx.x%) | xx (xx.x%) | xx (xx.x%) |
| Other | xx (xx.x%) | xx (xx.x%) | xx (xx.x%) |
| **Education** | | | |
| None | xx (xx.x%) | xx (xx.x%) | xx (xx.x%) |
| Primary school | xx (xx.x%) | xx (xx.x%) | xx (xx.x%) |
| Secondary school | xx (xx.x%) | xx (xx.x%) | xx (xx.x%) |
| Undergraduate degree | xx (xx.x%) | xx (xx.x%) | xx (xx.x%) |
| Postgraduate degree | xx (xx.x%) | xx (xx.x%) | xx (xx.x%) |
| Other technical / vocational training | xx (xx.x%) | xx (xx.x%) | xx (xx.x%) |
| Other | xx (xx.x%) | xx (xx.x%) | xx (xx.x%) |
| **Occupation** | | | |
| Management | xx (xx.x%) | xx (xx.x%) | xx (xx.x%) |
| Professional and related | xx (xx.x%) | xx (xx.x%) | xx (xx.x%) |
| Service | xx (xx.x%) | xx (xx.x%) | xx (xx.x%) |

Docusign Envelope ID: CD9CCBFF-A0AC-4C1B-A386-8B23A4692D4E

| | | | |
|---|---|---|---|
| Sales / Commercial | xx (xx.x%) | xx (xx.x%) | xx (xx.x%) |
| Construction | xx (xx.x%) | xx (xx.x%) | xx (xx.x%) |
| Armed forces | xx (xx.x%) | xx (xx.x%) | xx (xx.x%) |
| Farming/ Forestry/ fishing and related | xx (xx.x%) | xx (xx.x%) | xx (xx.x%) |
| Clerical/ administrative support | xx (xx.x%) | xx (xx.x%) | xx (xx.x%) |
| Installation and related | xx (xx.x%) | xx (xx.x%) | xx (xx.x%) |
| Manufacture and production | xx (xx.x%) | xx (xx.x%) | xx (xx.x%) |
| Transportation/ driver | xx (xx.x%) | xx (xx.x%) | xx (xx.x%) |
| No lifetime occupation | xx (xx.x%) | xx (xx.x%) | xx (xx.x%) |
| **Employment** | | | |
| Full-time paid work | xx (xx.x%) | xx (xx.x%) | xx (xx.x%) |
| Part-time paid work | xx (xx.x%) | xx (xx.x%) | xx (xx.x%) |
| Retired | xx (xx.x%) | xx (xx.x%) | xx (xx.x%) |
| Unemployed / redundant | xx (xx.x%) | xx (xx.x%) | xx (xx.x%) |
| Home duties | xx (xx.x%) | xx (xx.x%) | xx (xx.x%) |
| Student | xx (xx.x%) | xx (xx.x%) | xx (xx.x%) |
| Other | xx (xx.x%) | xx (xx.x%) | xx (xx.x%) |
| **Total number of people who usually live in household with participant** | | | |
| N Mean (SD) | N xx.x (xx.x) | N xx.x (xx.x) | N xx.x (xx.x) |
| **Smoking status** | | | |
| Non-smoker | xx (xx.x%) | xx (xx.x%) | xx (xx.x%) |
| Ex-smoker | xx (xx.x%) | xx (xx.x%) | xx (xx.x%) |

Docusign Envelope ID: CD9CCBFF-A0AC-4C1B-A386-8B23A4692D4E

| | | | |
|---|---|---|---|
| Time since stopped | N xx.x (xx.x) | N xx.x (xx.x) | N xx.x (xx.x) |
| Number of pack years(2) | N xx.x (xx.x) | N xx.x (xx.x) | N xx.x (xx.x) |
| Current smoker | xx (xx.x%) | xx (xx.x%) | xx (xx.x%) |
| Number of pack years(2) | N xx.x (xx.x) | N xx.x (xx.x) | N xx.x (xx.x) |
| **Location of ICH** | | | |
| Cortical | xx (xx.x%) | xx (xx.x%) | xx (xx.x%) |
| Deep | xx (xx.x%) | xx (xx.x%) | xx (xx.x%) |
| **Presumed etiology of ICH** | | | |
| Hypertension | xx (xx.x%) | xx (xx.x%) | xx (xx.x%) |
| Anticoagulation-related, e.g. warfarin-related | xx (xx.x%) | xx (xx.x%) | xx (xx.x%) |
| Antiplatelet-related, e.g. aspirin-related | xx (xx.x%) | xx (xx.x%) | xx (xx.x%) |
| Cerebral amyloid angiopathy related | xx (xx.x%) | xx (xx.x%) | xx (xx.x%) |
| **Dietary intake** | | | |
| **How many days per week do you eat fresh fruit** | | | |
| Never | xx (xx.x%) | xx (xx.x%) | xx (xx.x%) |
| < 1 per week | xx (xx.x%) | xx (xx.x%) | xx (xx.x%) |
| 1-2 per week | xx (xx.x%) | xx (xx.x%) | xx (xx.x%) |
| 3-4 per week | xx (xx.x%) | xx (xx.x%) | xx (xx.x%) |
| 5-6 per week | xx (xx.x%) | xx (xx.x%) | xx (xx.x%) |
| Everyday | xx (xx.x%) | xx (xx.x%) | xx (xx.x%) |
| **How many days per week do you eat uncooked vegetables** | | | |
| Never | xx (xx.x%) | xx (xx.x%) | xx (xx.x%) |

| | | | |
|---|---|---|---|
| < 1 per week | xx (xx.x%) | xx (xx.x%) | xx (xx.x%) |
| 1-2 per week | xx (xx.x%) | xx (xx.x%) | xx (xx.x%) |
| 3-4 per week | xx (xx.x%) | xx (xx.x%) | xx (xx.x%) |
| 5-6 per week | xx (xx.x%) | xx (xx.x%) | xx (xx.x%) |
| Everyday | xx (xx.x%) | xx (xx.x%) | xx (xx.x%) |
| **How many days per week do you eat cooked vegetables** | | | |
| Never | xx (xx.x%) | xx (xx.x%) | xx (xx.x%) |
| < 1 per week | xx (xx.x%) | xx (xx.x%) | xx (xx.x%) |
| 1-2 per week | xx (xx.x%) | xx (xx.x%) | xx (xx.x%) |
| 3-4 per week | xx (xx.x%) | xx (xx.x%) | xx (xx.x%) |
| 5-6 per week | xx (xx.x%) | xx (xx.x%) | xx (xx.x%) |
| Everyday | xx (xx.x%) | xx (xx.x%) | xx (xx.x%) |
| **How many days per week do you eat fish** | | | |
| Never | xx (xx.x%) | xx (xx.x%) | xx (xx.x%) |
| < 1 per week | xx (xx.x%) | xx (xx.x%) | xx (xx.x%) |
| 1-2 per week | xx (xx.x%) | xx (xx.x%) | xx (xx.x%) |
| 3-4 per week | xx (xx.x%) | xx (xx.x%) | xx (xx.x%) |
| 5-6 per week | xx (xx.x%) | xx (xx.x%) | xx (xx.x%) |
| Everyday | xx (xx.x%) | xx (xx.x%) | xx (xx.x%) |
| **Alcohol consumption (once per week or more)** | xx (xx.x%) | xx (xx.x%) | xx (xx.x%) |
| Average standard drinks (beer/wine/spirit) per week | N xx.x (xx.x) | N xx.x (xx.x) | N xx.x (xx.x) |
| Caffeinated drinks consumption currently | xx (xx.x%) | xx (xx.x%) | xx (xx.x%) |

Docusign Envelope ID: CD9CCBFF-A0AC-4C1B-A386-8B23A4692D4E

| | | | |
|---|---|---|---|
| incl. coffee, tea and other caffeinated drinks | | | |
| Average number of caffeinated drinks per week | N xx.x (xx.x) | N xx.x (xx.x) | N xx.x (xx.x) |

1. *East Asian includes Chinese, Japanese, Korean, Taiwanese and Vietnamese; South Asian includes Indian, Pakistani, Sri Lankan and Bangladeshi; South East Asian includes Indonesian, Malaysian and Filipino.*

2. *One pack of cigarettes per day for 5 years equals to 5 pack years.*

**Table 5. Medical History**

| Medical History | Intervention | Control | Total |
|---|---|---|---|
| Ischaemic stroke | xx (xx.x%) | xx (xx.x%) | xx (xx.x%) |
| ICH (i.e. prior to qualifying ICH) | xx (xx.x%) | xx (xx.x%) | xx (xx.x%) |
| Stroke of unknown type | xx (xx.x%) | xx (xx.x%) | xx (xx.x%) |
| Transient ischemic attack | xx (xx.x%) | xx (xx.x%) | xx (xx.x%) |
| Ischemic heart disease / coronary artery disease | xx (xx.x%) | xx (xx.x%) | xx (xx.x%) |
| Myocardial infarction | xx (xx.x%) | xx (xx.x%) | xx (xx.x%) |
| Coronary bypass grafting or coronary intervention | xx (xx.x%) | xx (xx.x%) | xx (xx.x%) |
| Valvular heart disease | xx (xx.x%) | xx (xx.x%) | xx (xx.x%) |
| Atrial fibrillation | xx (xx.x%) | xx (xx.x%) | xx (xx.x%) |
| Peripheral arterial disease | xx (xx.x%) | xx (xx.x%) | xx (xx.x%) |
| Hypertension | xx (xx.x%) | xx (xx.x%) | xx (xx.x%) |
| Depression requiring treatment | xx (xx.x%) | xx (xx.x%) | xx (xx.x%) |
| Chronic kidney disease | xx (xx.x%) | xx (xx.x%) | xx (xx.x%) |
| Diabetes mellitus Type I | xx (xx.x%) | xx (xx.x%) | xx (xx.x%) |
| Diabetes mellitus Type II | xx (xx.x%) | xx (xx.x%) | xx (xx.x%) |
| Falls | xx (xx.x%) | xx (xx.x%) | xx (xx.x%) |
| Number of falls in the past year | N Median (Q1-Q3) | N Median (Q1-Q3) | N Median (Q1-Q3) |
| Migraine | xx (xx.x%) | xx (xx.x%) | xx (xx.x%) |
| Time since onset (years) | N Mean(SE) | N Mean(SE) | N Mean(SE) |

Docusign Envelope ID: CD9CCBFF-A0AC-4C1B-A386-8B23A4692D4E

#### Table 6. Laboratory measurements

| Plasma Biochemistry - Fasting | Intervention<br>N Mean (SD) | Control<br>N Mean (SD) | Total<br>N Mean (SD) |
|---|---|---|---|
| **Glucose (mmol/l)** | | | |
| Repeat run-in/Baseline | N xx.x (xx.x) | N xx.x (xx.x) | N xx.x (xx.x) |
| Month xx (EOS) | N xx.x (xx.x) | N xx.x (xx.x) | N xx.x (xx.x) |
| **Total cholesterol (mmol/l)** | | | |
| Repeat run-in/Baseline | N xx.x (xx.x) | N xx.x (xx.x) | N xx.x (xx.x) |
| Month xx /EOS | N xx.x (xx.x) | N xx.x (xx.x) | N xx.x (xx.x) |
| **LDL (mmol/l)** | | | |
| Repeat run-in/Baseline | N xx.x (xx.x) | N xx.x (xx.x) | N xx.x (xx.x) |
| Month xx | N xx.x (xx.x) | N xx.x (xx.x) | N xx.x (xx.x) |
| **HDL (mmol/l)** | | | |
| Repeat run-in/Baseline | N xx.x (xx.x) | N xx.x (xx.x) | N xx.x (xx.x) |
| Month xx | N xx.x (xx.x) | N xx.x (xx.x) | N xx.x (xx.x) |
| **Triglycerides (mmol/l)** | | | |
| Repeat run-in/Baseline | N xx.x (xx.x) | N xx.x (xx.x) | N xx.x (xx.x) |
| Month xx | N xx.x (xx.x) | N xx.x (xx.x) | N xx.x (xx.x) |
| **Glucose (mmol/l)** | | | |
| Repeat run-in/Baseline | N xx.x (xx.x) | N xx.x (xx.x) | N xx.x (xx.x) |
| Month xx | N xx.x (xx.x) | N xx.x (xx.x) | N xx.x (xx.x) |
| **Creatinine (mmol/l)** | | | |
| Repeat run-in/Baseline | N xx.x (xx.x) | N xx.x (xx.x) | N xx.x (xx.x) |
| Week 6 | N xx.x (xx.x) | N xx.x (xx.x) | N xx.x (xx.x) |
| Month xx | N xx.x (xx.x) | N xx.x (xx.x) | N xx.x (xx.x) |
| **Sodium (mmol/l)** | | | |
| Repeat run-in/Baseline | N xx.x (xx.x) | N xx.x (xx.x) | N xx.x (xx.x) |
| Week 6 | N xx.x (xx.x) | N xx.x (xx.x) | N xx.x (xx.x) |
| Month xx (EOS) | N xx.x (xx.x) | N xx.x (xx.x) | N xx.x (xx.x) |
| **Potassium (mmol/l)** | | | |
| Repeat run-in/Baseline | N xx.x (xx.x) | N xx.x (xx.x) | N xx.x (xx.x) |
| Week 6 | N xx.x (xx.x) | N xx.x (xx.x) | N xx.x (xx.x) |
| Month xx (EOS) | N xx.x (xx.x) | N xx.x (xx.x) | N xx.x (xx.x) |

Docusign Envelope ID: CD9CCBFF-A0AC-4C1B-A386-8B23A4692D4E

| | | | |
|---|---|---|---|
| **Blood urea nitrogen (mmol/l)** | | | |
| Repeat run-in/Baseline | N xx.x (xx.x) | N xx.x (xx.x) | N xx.x (xx.x) |
| Week 6 | N xx.x (xx.x) | N xx.x (xx.x) | N xx.x (xx.x) |
| Month xx (EOS) | N xx.x (xx.x) | N xx.x (xx.x) | N xx.x (xx.x) |
| **eGFR (CKD-EPI)** | | | |
| Repeat run-in/Baseline | N xx.x (xx.x) | N xx.x (xx.x) | N xx.x (xx.x) |
| Week 6 | N xx.x (xx.x) | N xx.x (xx.x) | N xx.x (xx.x) |
| Month xx (EOS) | N xx.x (xx.x) | N xx.x (xx.x) | N xx.x (xx.x) |
| **eGFR >90 mL/min/1.73m$^2$ (CKD-EPI)** | | | |
| Repeat run-in/Baseline | N=xxx | N=xxx | N=xxx |
| Week 6 | n (xx.x%) | n (xx.x%) | n (xx.x%) |
| Month xx (EOS) | n (xx.x%) | n (xx.x%) | n (xx.x%) |
| **Aspartate aminotransferase AST (IU/l)** | | | |
| Repeat run-in/Baseline | N xx.x (xx.x) | N xx.x (xx.x) | N xx.x (xx.x) |
| Month xx (EOS) | N xx.x (xx.x) | N xx.x (xx.x) | N xx.x (xx.x) |
| **Alanine aminotransferase ALT (IU/l)** | | | |
| Repeat run-in/Baseline | N xx.x (xx.x) | N xx.x (xx.x) | N xx.x (xx.x) |
| Month xx (EOS) | N xx.x (xx.x) | N xx.x (xx.x) | N xx.x (xx.x) |

Docusign Envelope ID: CD9CCBFF-A0AC-4C1B-A386-8B23A4692D4E

#### Table 7. Blood Pressure / Clinical assessment

| | Intervention | | Control | |
|---|---|---|---|---|
| | Actual values n/ Mean (SE) | Change from baseline Mean (SE) | Actual values n/ Mean (SE) | Change from baseline Mean (SE) |
| **SBP (mmHg)** | | | | |
| Randomisation* | n/xx.xx (xx.x) | | n/xx.xx (xx.x) | |
| 6 Weeks | n/xx.xx (xx.x) | xx.xx (xx.x) | n/xx.xx (xx.x) | xx.xx (xx.x) |
| 6 Months | n/xx.xx (xx.x) | xx.xx (xx.x) | n/xx.xx (xx.x) | xx.xx (xx.x) |
| 12 Months | n/xx.xx (xx.x) | xx.xx (xx.x) | n/xx.xx (xx.x) | xx.xx (xx.x) |
| 24 Months | n/xx.xx (xx.x) | xx.xx (xx.x) | n/xx.xx (xx.x) | xx.xx (xx.x) |
| 36 Months | n/xx.xx (xx.x) | xx.xx (xx.x) | n/xx.xx (xx.x) | xx.xx (xx.x) |
| Repeated for all timepoints | | | | |
| **DBP (mmHg)** | | | | |
| Randomisation* | n/xx.xx (xx.x) | | n/xx.xx (xx.x) | |
| 6 Weeks | n/xx.xx (xx.x) | xx.xx (xx.x) | n/xx.xx (xx.x) | xx.xx (xx.x) |
| 6 Months | n/xx.xx (xx.x) | xx.xx (xx.x) | n/xx.xx (xx.x) | xx.xx (xx.x) |
| 12 Months | n/xx.xx (xx.x) | xx.xx (xx.x) | n/xx.xx (xx.x) | xx.xx (xx.x) |
| 24 Months | n/xx.xx (xx.x) | xx.xx (xx.x) | n/xx.xx (xx.x) | xx.xx (xx.x) |
| 36 Months | n/xx.xx (xx.x) | xx.xx (xx.x) | n/xx.xx (xx.x) | xx.xx (xx.x) |
| Repeated for all timepoints | | | | |
| **Heart rate (bpm)** | | | | |
| Randomisation | n/xx.xx (xx.x) | | n/xx.xx (xx.x) | |
| 6 Weeks | n/xx.xx (xx.x) | xx.xx (xx.x) | n/xx.xx (xx.x) | xx.xx (xx.x) |
| 6 Months | n/xx.xx (xx.x) | xx.xx (xx.x) | n/xx.xx (xx.x) | xx.xx (xx.x) |
| 12 Months | n/xx.xx (xx.x) | xx.xx (xx.x) | n/xx.xx (xx.x) | xx.xx (xx.x) |
| 24 Months | n/xx.xx (xx.x) | xx.xx (xx.x) | n/xx.xx (xx.x) | xx.xx (xx.x) |
| Repeated for all timepoints | | | | |

**BP measurements at randomisation refer to average of 2 measurements taken from the latest assessment (run-in or re-run visits).*

Docusign Envelope ID: CD9CCBFF-A0AC-4C1B-A386-8B23A4692D4E

**Table 8. Between group differences in Blood Pressure before and after drug formulation switch**

| | Last visit of Triple pill therapy | | | First visit after switch to GMRx2 | | |
|---|---|---|---|---|---|---|
| | Intervention Mean (SE) | Control Mean (SE) | MD (95%CI) | Intervention Mean (SE) | Control Mean (SE) | MD (95%CI) |
| SBP (mmHg) | xx.xx (xx.x) | xx.xx (xx.x) | xx.xx (xx.x, xx.x) | xx.xx (xx.x) | xx.xx (xx.x) | xx.xx (xx.x, xx.x) |
| DBP (mmHg) | xx.xx (xx.x) | xx.xx (xx.x) | xx.xx (xx.x, xx.x) | xx.xx (xx.x) | xx.xx (xx.x) | xx.xx (xx.x, xx.x) |

MD=mean difference

Docusign Envelope ID: CD9CCBFF-A0AC-4C1B-A386-8B23A4692D4E

**Table 9. smRS descriptive table by timepoints**

| | Intervention | Control | Total |
|---|---|---|---|
| **Baseline** | **N=xxx** | **N=xxx** | **N=xxx** |
| **0** | xx (xx.x%) | xx (xx.x%) | xx (xx.x%) |
| **1** | xx (xx.x%) | xx (xx.x%) | xx (xx.x%) |
| **2** | xx (xx.x%) | xx (xx.x%) | xx (xx.x%) |
| **3** | xx (xx.x%) | xx (xx.x%) | xx (xx.x%) |
| **4** | xx (xx.x%) | xx (xx.x%) | xx (xx.x%) |
| **5** | xx (xx.x%) | xx (xx.x%) | xx (xx.x%) |
| **6** | xx (xx.x%) | xx (xx.x%) | xx (xx.x%) |
| Independent [mRS 0-2] | xx (xx.x%) | xx (xx.x%) | xx (xx.x%) |
| Dependent [mRS 3-6] | xx (xx.x%) | xx (xx.x%) | xx (xx.x%) |
| **12 Months** | **N=xxx** | **N=xxx** | **N=xxx** |
| **0** | xx (xx.x%) | xx (xx.x%) | xx (xx.x%) |
| **1** | xx (xx.x%) | xx (xx.x%) | xx (xx.x%) |
| **2** | xx (xx.x%) | xx (xx.x%) | xx (xx.x%) |
| **3** | xx (xx.x%) | xx (xx.x%) | xx (xx.x%) |
| **4** | xx (xx.x%) | xx (xx.x%) | xx (xx.x%) |
| **5** | xx (xx.x%) | xx (xx.x%) | xx (xx.x%) |
| **6** | xx (xx.x%) | xx (xx.x%) | xx (xx.x%) |
| Independent [mRS 0-2] | xx (xx.x%) | xx (xx.x%) | xx (xx.x%) |
| Dependent [mRS 3-6] | xx (xx.x%) | xx (xx.x%) | xx (xx.x%) |
| **24 Months** | **N=xxx** | **N=xxx** | **N=xxx** |
| **0** | xx (xx.x%) | xx (xx.x%) | xx (xx.x%) |
| **1** | xx (xx.x%) | xx (xx.x%) | xx (xx.x%) |
| **2** | xx (xx.x%) | xx (xx.x%) | xx (xx.x%) |
| **3** | xx (xx.x%) | xx (xx.x%) | xx (xx.x%) |
| **4** | xx (xx.x%) | xx (xx.x%) | xx (xx.x%) |
| **5** | xx (xx.x%) | xx (xx.x%) | xx (xx.x%) |
| **6** | xx (xx.x%) | xx (xx.x%) | xx (xx.x%) |
| Independent [mRS 0-2] | xx (xx.x%) | xx (xx.x%) | xx (xx.x%) |

Docusign Envelope ID: CD9CCBFF-A0AC-4C1B-A386-8B23A4692D4E

| | | | |
|---|---|---|---|
| Dependent [mRS 3-6] | xx (xx.x%) | xx (xx.x%) | xx (xx.x%) |
| **36 Months** | **N=xxx** | **N=xxx** | **N=xxx** |
| **0** | xx (xx.x%) | xx (xx.x%) | xx (xx.x%) |
| **1** | xx (xx.x%) | xx (xx.x%) | xx (xx.x%) |
| **2** | xx (xx.x%) | xx (xx.x%) | xx (xx.x%) |
| **3** | xx (xx.x%) | xx (xx.x%) | xx (xx.x%) |
| **4** | xx (xx.x%) | xx (xx.x%) | xx (xx.x%) |
| **5** | xx (xx.x%) | xx (xx.x%) | xx (xx.x%) |
| **6** | xx (xx.x%) | xx (xx.x%) | xx (xx.x%) |
| Independent [mRS 0-2] | xx (xx.x%) | xx (xx.x%) | xx (xx.x%) |
| Dependent [mRS 3-6] | xx (xx.x%) | xx (xx.x%) | xx (xx.x%) |

*Repeated for month 48, 60, 72 and EOS*

Docusign Envelope ID: CD9CCBFF-A0AC-4C1B-A386-8B23A4692D4E

#### Table 10. EQ5D-3L – descriptive table by timepoints

| EQ5D-3L | Intervention | Control | Total |
|---|---|---|---|
| **Baseline** | **N=xxx** | **N=xxx** | **N=xxx** |
| Overall self-reported health score (VAS) - mean(SD) | xx.xx (xx.x) | xx.xx (xx.x) | xx.xx (xx.x) |
| Health utility score – mean_(SD) | xx.xx (xx.x) | xx.xx (xx.x) | xx.xx (xx.x) |
| **12 Months** | **N=xxx** | **N=xxx** | **N=xxx** |
| Overall self-reported health score (VAS) - mean(SD) | xx.xx (xx.x) | xx.xx (xx.x) | xx.xx (xx.x) |
| Health utility score – mean_(SD) | xx.xx (xx.x) | xx.xx (xx.x) | xx.xx (xx.x) |
| **Repeat for month 24, 36, 48, 60, 72, EOS** | | | |

Docusign Envelope ID: CD9CCBFF-A0AC-4C1B-A386-8B23A4692D4E

**Table 11. MoCA - descriptive table by timepoints**

| MoCA | Intervention | Control | Total |
|---|---|---|---|
| Baseline | N=xxx | N=xxx | N=xxx |
| Total score | x.xx (x.xx) | x.xx (x.xx) | x.xx (x.x) |
| 6 months | N=xxx | N=xxx | N=xxx |
| Total score | x.xx (x.xx) | x.xx (x.xx) | x.xx (x.x) |
| Repeat at 18, 30, 42, 54, 66 months & EOS | | | |

Docusign Envelope ID: CD9CCBFF-A0AC-4C1B-A386-8B23A4692D4E

**Table 12. BMET - descriptive table by timepoints**

| BMET | Intervention | Control | Total |
|---|---|---|---|
| **Baseline** | **N=xxx** | **N=xxx** | **N=xxx** |
| Orientation (score) | x.xx (x.xx) | x.xx (x.xx) | x.xx (x.xx) |
| Five item repetition (score) | x.xx (x.xx) | x.xx (x.xx) | x.xx (x.xx) |
| Letter-Number matching (score) | x.xx (x.xx) | x.xx (x.xx) | x.xx (x.xx) |
| Motor sequencing (seconds) | x.xx (x.xx) | x.xx (x.xx) | x.xx (x.xx) |
| Letter sequencing (seconds) | x.xx (x.xx) | x.xx (x.xx) | x.xx (x.xx) |
| Number-Letter sequencing (seconds) | x.xx (x.xx) | x.xx (x.xx) | x.xx (x.xx) |
| Five item memory (delayed recall) (score) | x.xx (x.xx) | x.xx (x.xx) | x.xx (x.xx) |
| Five item memory (delayed recognition) (score) | x.xx (x.xx) | x.xx (x.xx) | x.xx (x.xx) |
| Aggregated score | | | |
| **6 months** | **N=xxx** | **N=xxx** | **N=xxx** |
| Orientation (score) | x.xx (x.xx) | x.xx (x.xx) | x.xx (x.xx) |
| Five item repetition (score) | x.xx (x.xx) | x.xx (x.xx) | x.xx (x.xx) |
| Letter-Number matching (score) | x.xx (x.xx) | x.xx (x.xx) | x.xx (x.xx) |
| Motor sequencing (seconds) | x.xx (x.xx) | x.xx (x.xx) | x.xx (x.xx) |
| Letter sequencing (seconds) | x.xx (x.xx) | x.xx (x.xx) | x.xx (x.xx) |
| Number-Letter sequencing (seconds) | x.xx (x.xx) | x.xx (x.xx) | x.xx (x.xx) |
| Five item memory (delayed recall) (score) | x.xx (x.xx) | x.xx (x.xx) | x.xx (x.xx) |
| Five item memory (delayed recognition) (score) | x.xx (x.xx) | x.xx (x.xx) | x.xx (x.xx) |
| Aggregated score | | | |
| **Repeat at 18, 30, 42, 54, 66 months & EOS** | | | |

Docusign Envelope ID: CD9CCBFF-A0AC-4C1B-A386-8B23A4692D4E

**Table 13. Primary secondary and exploratory outcomes – model results**

| Outcomes / Analysis method | Raw estimate | | Model result[1] | | |
|---|---|---|---|---|---|
| | Intervention n/N (%) / mean (SD) | Control n/N (%) / mean (SD) | OR/MD/HR (95% CI) | p-value | Holm-Sidak adjustment |
| **Primary outcome** | | | | | n/a |
| Time to first occurrence of recurrent stroke (ICH/ischaemic/undifferentiated) | n/N (%) | n/N (%) | | | |
| Cause-specific hazard model – unadjusted[1] (survival) | | | xx.xx (xx.xx to xx.xx) | x.xxx | |
| Cause-specific hazard model – adjusted[2] (survival) | | | xx.xx (xx.xx to xx.xx) | x.xxx | |
| Fine and gray model – unadjusted[1] (survival) | | | xx.xx (xx.xx to xx.xx) | x.xxx | |
| **Secondary efficacy outcomes** | | | | | |
| Time to MACE (CV death, non-fatal MI, or non-fatal stroke) (survival) | n/N (%) | n/N (%) | xx.xx (xx.xx to xx.xx) | x.xxx | x.xxx |
| Time to cardiovascular mortality (survival) | n/N (%) | n/N (%) | xx.xx (xx.xx to xx.xx) | x.xxx | x.xxx |
| Hypertension control (SBP<130 mmHg) at 6 months (binary) | n/N (%) | n/N (%) | xx.xx (xx.xx to xx.xx) | x.xxx | x.xxx |

Docusign Envelope ID: CD9CCBFF-A0AC-4C1B-A386-8B23A4692D4E

| | | | | | |
|---|---|---|---|---|---|
| **Other outcomes** | | | | | n/a |
| **Other cardiovascular and mortality outcomes** | | | | | |
| Time to first recurrence of ICH (survival) | n/N (%) | n/N (%) | xx.xx (xx.xx to xx.xx) | x.xxx | |
| Time to first occurrence of ischemic stroke (survival) | n/N (%) | n/N (%) | xx.xx (xx.xx to xx.xx) | x.xxx | |
| Time to first occurrence of fatal stroke (survival) | n/N (%) | n/N (%) | xx.xx (xx.xx to xx.xx) | x.xxx | |
| Time to first occurrence of stroke of undifferentiated origin (survival) | n/N (%) | n/N (%) | xx.xx (xx.xx to xx.xx) | x.xxx | |
| Time to all-cause mortality | n/N (%) | n/N (%) | xx.xx (xx.xx to xx.xx) | x.xxx | |
| **Cognitive outcomes** | | | | | |
| Dementia-free survival (binomial) [3] | Annual event rate (95% C.I) | Annual event rate (95% C.I) | IRR<br>xx.xx(xx.xx to xx.xx) | x.xxx | |
| Sensitivity analysis | | | | | |
| Dementia-free survival adjusted for education level (>12 years vs. other) (binomial)[3] | Annual event rate (95% C.I) | Annual event rate (95% C.I) | IRR<br>xx.xx(xx.xx to xx.xx) | x.xxx | |

| | | | | |
|---|---|---|---|---|
| Dementia-free survival with missing values imputed as dementia cases (binomial) [3] | Annual event rate (95% C.I) | Annual event rate (95% C.I) | IRR xx.xx(xx.xx to xx.xx) | x.xxx |
| Tipping point analysis[3] | Annual event rate (95% C.I) | Annual event rate (95% C.I) | IRR xx.xx(xx.xx to xx.xx) | x.xxx |
| Alive and free of MCI (binomial) [3] | Annual event rate (95% C.I) | Annual event rate (95% C.I) | IRR xx.xx(xx.xx to xx.xx) | x.xxx |
| Alive and free of MCI with missing values imputed as MCI cases sensitivity analysis (binomial) [3] | Annual event rate (95% C.I) | Annual event rate (95% C.I) | IRR xx.xx(xx.xx to xx.xx) | x.xxx |
| Composite of free and alive of dementia and MCI diagnosis (binomial) [3] | Annual event rate (95% C.I) | Annual event rate (95% C.I) | IRR xx.xx(xx.xx to xx.xx) | x.xxx |
| Cognitive decline MoCA (continuous) | n/N (%) | n/N (%) | xx.xx (xx.xx to xx.xx) | x.xxx |
| **Disability outcomes** | | | | |
| Disability-free survival (death or dependency on the mRS [0-2 vs. 3-6]) (binomial) [3] | n/N (%) | n/N (%) | xx.xx (xx.xx to xx.xx) | x.xxx |
| Health utility score EQ5D-3L (continuous) | Mean (SD) | Mean (SD) | xx.xx (xx.xx to xx.xx) | x.xxx |

Docusign Envelope ID: CD9CCBFF-A0AC-4C1B-A386-8B23A4692D4E

| | | | | | |
|---|---|---|---|---|---|
| Physical functioning smRS score- (ordinal) | | | xx.xx (xx.xx to xx.xx) | x.xxx | |
| **Other blood pressure outcomes** | | | | | |
| Hypertension control (SBP<130 mmHg) at final follow-up (binomial) [3] | Annual event rate (95% C.I) | Annual event rate (95% C.I) | IRR<br>xx.xx(xx.xx to xx.xx) | x.xxx | |
| Change in SBP from randomization (cont.) | Mean (SD) | Mean (SD) | xx.xx (xx.xx to xx.xx) | x.xxx | |
| Change in DBP from randomization (cont.) | Mean (SD) | Mean (SD) | xx.xx (xx.xx to xx.xx) | x.xxx | |
| Time at target (SBP<130mmHg throughout follow-up) (cont.) | Mean (SD) | Mean (SD) | xx.xx (xx.xx to xx.xx) | x.xxx | |
| **Other exploratory outcomes** | | | | | |
| BMET score (continuous) | Mean (SD) | Mean (SD) | xx.xx (xx.xx to xx.xx) | x.xxx | n/a |

1. *All analyses have been adjusted for the stratification variables of country of recruitment, age (<65 vs. ≥65 years) and baseline SBP (<140 vs ≥140mmHg) as fixed effects. For hypertension related outcomes, BP Ordinal, binary and continuous models consist of generalised linear mixed models with appropriate distribution and link function.*
2. *Adjusted models include the following additional baseline covariates: sex, pre-morbid level of function on the smRS, history of hypertension, history of cardiac disease, history of diabetes mellitis, level of education (>=12 years or other)*
3. *Binomial regression adjusted for log of follow-up duration (years between randomization and end of follow-up) using an offset. We will thus derive a yearly incidence rate of death or dementia which will be compared between treatment arms using an incidence rate ratio and 95% CI.*

Docusign Envelope ID: CD9CCBFF-A0AC-4C1B-A386-8B23A4692D4E

**Table 14. Win Ratio analysis**

| Outcome | Intervention wins (N %) | Control wins (N %) | Ties (N%) | Win ratio (95% CI) | p-value | Win odds (95% CI) |
|---|---|---|---|---|---|---|
| Primary composite | xxx 0.xxx% | xxx 0.xxx% | xxx 0.xxx% | xx.xx (xx.xx to xx.xx) | 0.xxx | xx.xx (xx.xx to xx.xx) |
| Time to CV death | xxx 0.xxx% | xxx 0.xxx% | xxx 0.xxx% | xx.xx (xx.xx to xx.xx) | 0.xxx | xx.xx (xx.xx to xx.xx) |
| Time to non-CV death | xxx 0.xxx% | xxx 0.xxx% | xxx 0.xxx% | xx.xx (xx.xx to xx.xx) | 0.xxx | xx.xx (xx.xx to xx.xx) |
| Time to Nonfatal stroke | xxx 0.xxx% | xxx 0.xxx% | xxx 0.xxx% | xx.xx (xx.xx to xx.xx) | 0.xxx | xx.xx (xx.xx to xx.xx) |
| Dementia | xxx 0.xxx% | xxx 0.xxx% | xxx 0.xxx% | xx.xx (xx.xx to xx.xx) | 0.xxx | xx.xx (xx.xx to xx.xx) |
| Time to Nonfatal MI | xxx 0.xxx% | xxx 0.xxx% | xxx 0.xxx% | xx.xx (xx.xx to xx.xx) | 0.xxx | xx.xx (xx.xx to xx.xx) |
| Dependency [mRS ≥3] | xxx 0.xxx% | xxx 0.xxx% | xxx 0.xxx% | xx.xx (xx.xx to xx.xx) | 0.xxx | xx.xx (xx.xx to xx.xx) |
| MCI | xxx 0.xxx% | xxx 0.xxx% | xxx 0.xxx% | xx.xx (xx.xx to xx.xx) | 0.xxx | xx.xx (xx.xx to xx.xx) |

** For or time-to-event outcomes (e.g. time to death or time to stroke), the comparison is only made on the shortest follow-up time in the pair. For example, if one patient from the pair was followed for 12 months and the other for 18 months, we will only determine the winner based on the first 12 months. This applies to steps 1, 2, 3 and 5.*

Docusign Envelope ID: CD9CCBFF-A0AC-4C1B-A386-8B23A4692D4E

#### Table 15. Compliance to study treatment over time

| | Intervention | Control | Total |
|---|---|---|---|
| **Week 6** | | | |
| **Number of days in which doses were missed in the last week as reported by participant (0 to 7)** | N=xxx | N=xxx | N=xxx |
| Mean (SD) | xx.x (xx.xx) | xx.x (xx.xx) | xx.x (xx.xx) |
| Median [Q1 - Q3] | xx (xx - xx) | xx (xx - xx) | xx (xx - xx) |
| Min, Max | xx, xx | xx, xx | xx, xx |
| **Has the patient brought in any remaining medication (yes/no)** | xxx (xx.x%) | xxx (xx.x%) | xxx (xx.x%) |
| **For participants in China:** | | | |
| **Number of capsules / pills remaining from returned medication** | N=xxx | N=xxx | N=xxx |
| Mean (SD) | xx.x (xx.xx) | xx.x (xx.xx) | xx.x (xx.xx) |
| Median [Q1 - Q3] | xx (xx - xx) | xx (xx - xx) | xx (xx - xx) |
| Min, Max | xx, xx | xx, xx | xx, xx |
| **Overall Adherence**<br><br>less than or equal to 15% (1/7 * 100 =14.3%)<br><br>based on self-reported missed doses (no more than 1 day missed out of 7 days) of study medication. | xx/xxx (xx.x%) | xx/xxx (xx.x%) | xxx (xx.x%) |
| ***Repeated for all follow-up timepoints*** | | | |

Docusign Envelope ID: CD9CCBFF-A0AC-4C1B-A386-8B23A4692D4E

#### Table 16. Protocol violations

| Protocol violations | Intervention | Control |
|---|---|---|
| Randomisation of ineligible patient | nEVT nPT (xx.x%) | nEVT nPT (xx.x%) |
| Incorrect Treatment Pack used | nEVT nPT (xx.x%) | nEVT nPT (xx.x%) |
| Prohibited or restricted medication use | nEVT nPT (xx.x%) | nEVT nPT (xx.x%) |
| Other | nEVT nPT (xx.x%) | nEVT nPT (xx.x%) |

Docusign Envelope ID: CD9CCBFF-A0AC-4C1B-A386-8B23A4692D4E

### Table 17. Concomitant medications

| | Intervention | Control | Total |
|---|---|---|---|
| **Baseline** | **N=xxx** | **N=xxx** | **N=xxx** |
| **Current BP lowering medication** | | | |
| Angiotensin converting enzyme inhibitor | xxx (xx.x%) | xxx (xx.x%) | xxx (xx.x%) |
| Angiotensin II receptor blockers | xxx (xx.x%) | xxx (xx.x%) | xxx (xx.x%) |
| Thiazide or thiazide-like diuretic | xxx (xx.x%) | xxx (xx.x%) | xxx (xx.x%) |
| Other diuretic | xxx (xx.x%) | xxx (xx.x%) | xxx (xx.x%) |
| Calcium channel blocker | xxx (xx.x%) | xxx (xx.x%) | xxx (xx.x%) |
| Beta blocker | xxx (xx.x%) | xxx (xx.x%) | xxx (xx.x%) |
| Other BP lowering medication | | | |
| **Week 6** | **N=xxx** | **N=xxx** | **N=xxx** |
| Agents acting on the renin-angiotensin system | xxx (xx.x%) | xxx (xx.x%) | xxx (xx.x%) |
| Antihypertensives | xxx (xx.x%) | xxx (xx.x%) | xxx (xx.x%) |
| Beta blocking agents | xxx (xx.x%) | xxx (xx.x%) | xxx (xx.x%) |
| Calcium channel blockers | xxx (xx.x%) | xxx (xx.x%) | xxx (xx.x%) |
| Cardiac therapy | xxx (xx.x%) | xxx (xx.x%) | xxx (xx.x%) |
| Diuretics | xxx (xx.x%) | xxx (xx.x%) | xxx (xx.x%) |
| Lipid modifying agents | xxx (xx.x%) | xxx (xx.x%) | xxx (xx.x%) |
| Vasoprotectives | xxx (xx.x%) | xxx (xx.x%) | xxx (xx.x%) |
| **Repeat for follow-up timepoints** | | | |

Docusign Envelope ID: CD9CCBFF-A0AC-4C1B-A386-8B23A4692D4E

**Table 18. Summary of adverse drug reactions**

| Adverse Events | Intervention<br><br>No. of events No. of patients (% ) | Control<br><br>No. of events No. of patients (% ) |
|---|---|---|
| **Adverse Drug Reactions** | nEVT nPT(xx.x%) | nEVT nPT(xx.x%) |
| **Adverse Drug Reactions resulting in treatment withdrawal** | nEVT nPT(xx.x%) | nEVT nPT(xx.x%) |
| **Serious Adverse Drug Reactions** | nEVT nPT(xx.x%) | nEVT nPT(xx.x%) |
| Resulted in death | nEVT nPT(xx.x%) | nEVT nPT(xx.x%) |
| Life threatening | nEVT nPT(xx.x%) | nEVT nPT(xx.x%) |
| Requires prolonged hospitalisation | nEVT nPT(xx.x%) | nEVT nPT(xx.x%) |
| Results in persistent or severe disability/incapacity | nEVT nPT(xx.x%) | nEVT nPT(xx.x%) |
| Results in congenital anomaly/birth defect | nEVT nPT(xx.x%) | nEVT nPT(xx.x%) |
| Is medically significant and may require intervention to prevent one of the above | nEVT nPT(xx.x%) | nEVT nPT(xx.x%) |
| **Suspected Unexpected Serious Adverse Reactions** | nEVT nPT(xx.x%) | nEVT nPT(xx.x%) |
| Resulted in death | nEVT nPT(xx.x%) | nEVT nPT(xx.x%) |
| Life threatening | nEVT nPT(xx.x%) | nEVT nPT(xx.x%) |
| Requires prolonged hospitalisation | nEVT nPT(xx.x%) | nEVT nPT(xx.x%) |
| Results in persistent or severe disability/incapacity | nEVT nPT(xx.x%) | nEVT nPT(xx.x%) |
| Results in congenital anomaly/birth defect | nEVT nPT(xx.x%) | nEVT nPT(xx.x%) |
| Is medically significant and may require intervention to prevent one of the above | nEVT nPT(xx.x%) | nEVT nPT(xx.x%) |

Docusign Envelope ID: CD9CCBFF-A0AC-4C1B-A386-8B23A4692D4E

| | | |
|---|---|---|
| **Adverse events of special interest (AESIs)** | nEVT nPT(xx.x%) | nEVT nPT(xx.x%) |
| Headache | nEVT nPT(xx.x%) | nEVT nPT(xx.x%) |
| Syncope/collapse | nEVT nPT(xx.x%) | nEVT nPT(xx.x%) |
| Falls | nEVT nPT(xx.x%) | nEVT nPT(xx.x%) |
| Pedal oedema/ankle swelling | nEVT nPT(xx.x%) | nEVT nPT(xx.x%) |
| Hypo/hyperkalaemia | nEVT nPT(xx.x%) | nEVT nPT(xx.x%) |
| Hyponatremia | nEVT nPT(xx.x%) | nEVT nPT(xx.x%) |

Docusign Envelope ID: CD9CCBFF-A0AC-4C1B-A386-8B23A4692D4E

#### Figure 2: Enrolment over time

#### Figure 3: Cumulative incidence function of time to occurrence of first recurrent stroke event

*Programming note: add number at risk every 6 months, display median and quartiles as well as results from the Cox model (hazard ratio, 95% CI and p-value)*

#### Figure 4. Kaplan-Meier plot of primary and stroke-related secondary outcomes

*Will show the p-value from the log-rank test and the number at risk in each treatment arm using a 6-month interval.*

#### Figure 5: Forest plot for subgroup analysis of time to first recurrent stroke event

#### Figure 6: Forest plot for stratified analysis of BP endpoints by IMP groups

Docusign Envelope ID: CD9CCBFF-A0AC-4C1B-A386-8B23A4692D4E

**Figure 7. Bar chart of smRS, MoCA and EQ5D-3L across all timepoints (Baseline and each follow-up timepoint) by treatment groups**

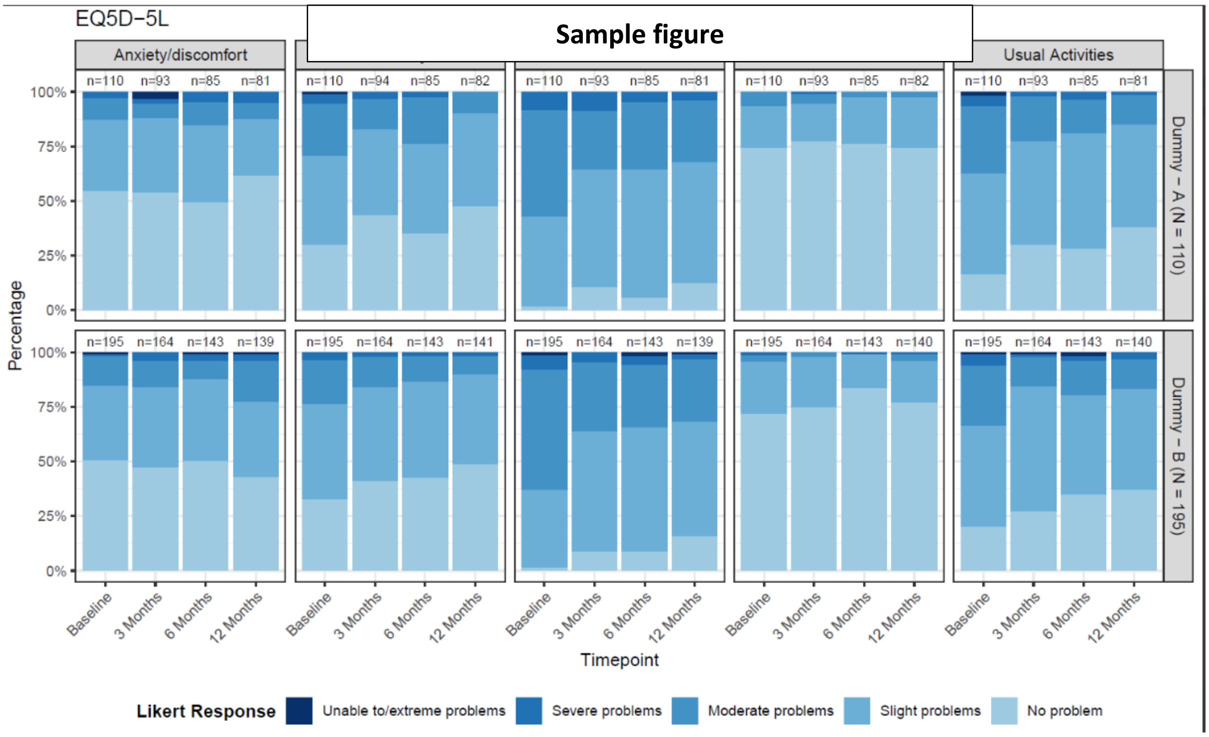

*Programming notes:*

- *Do this figure for all binary and ordinal variables including smRS, EQ5d-3L*
- *Create a stacked bar chart with two columns (one for each treatment group) for each timepoint including baseline and, within each column, stack the categories and colour code using a gradient (e.g. white to dark red).*
- *Display raw percentages within categories and denominators below the x-axis*

*Display overall odds ratio, 95% confidence interval and p-value obtained from a mixed logistic or proportional odds model (see Section 3.14.2,3.14.2.1 and 3.14.4 for details).*

Docusign Envelope ID: CD9CCBFF-A0AC-4C1B-A386-8B23A4692D4E

**Figure 8. Mean plot of Blood pressure, Health utility score, MoCA score and BMET aggregated score over time (Baseline to EOS) by intervention**

*Programming notes:*

- *Show means and 95%CI. Display raw means on the graph as numbers near each dot and denominators below the x-axis.*
- 
- *Also display overall mean difference, 95% confidence interval and p-value from repeated-measure linear mixed model across all follow-up timepoints (see Section xx for details)*

**Listing 1: SAE listing**

**Listing 2. AE listing**

**Listing 3. Protocol deviations**
